# Psychometric Properties of the ASEBA Child Behaviour Checklist and Youth Self-Report in Sub-Saharan Africa: A Systematic Review

**DOI:** 10.1101/2021.10.15.21265039

**Authors:** Michal R. Zieff, Claire Fourie, Michelle Hoogenhout, Kirsten A. Donald

## Abstract

**Objective:** Behavioural screening tools may be used to identify at-risk children in resource-limited settings in sub-Saharan Africa. The ASEBA forms (Child Behaviour Checklist and Youth Self-Report) are frequently translated and adapted for use in sub-Saharan African populations, but little is known about their measurement properties in these contexts.

**Methods:** We conducted a systematic review of all published journal articles that used the ASEBA forms with sub-Saharan African samples. We evaluated the reported psychometric properties, as well as the methodological quality of the psychometric evaluations, using COSMIN (COnsensus-based Standards for the selection of health Measurement INstruments) guidelines.

**Results:** Fifty-eight studies reported measurement properties of the ASEBA forms. Most studies came from Southern (*n* = 29, 50%) or East African (*n* = 25, 43%) countries. Forty-nine studies (84%) used translated versions of the tool, but details regarding the translation process, if available, were often sparse. Most studies (*n* = 47, 81%) only reported internal consistency (using coefficient alpha) for one or more subscales. The methodological quality of the psychometric evaluations ranged from ‘very good’ to ‘inadequate’ across all measurement properties, except for internal consistency.

**Conclusion:** There is limited good quality psychometric evidence available for the ASEBA forms in sub-Saharan Africa. We recommend (i) implementing a standardised procedure for conducting and reporting translation processes, and (ii) conducting more comprehensive psychometric evaluations of the translated versions of the tools.

---

Behavioural and emotional problems represent a significant health burden amongst children and adolescents in sub-Saharan Africa (SSA; M. A. Cortina et al., 2012; Kusi-Mensah et al., 2019; Mulatu, 1995; Ndetei et al., 2016). As specialist child mental health services in SSA are limited (Jenkins et al., 2010), screening tools, such as the Achenbach System of Empirically Based Assessment (ASEBA) forms (Achenbach & Rescorla, 2000, 2001), are often administered in community settings to detect common childhood emotional and behavioural problems (Hoosen et al., 2018). Screening tools typically do not require specialist training and are generally quick and easy to administer and score, making them particularly advantageous in low- and middle-income settings (Sharp et al., 2014). Despite their widespread use, there are no systematic reviews on the validity, reliability, and cultural appropriateness of the ASEBA forms in the diverse populations and contexts of SSA. This review seeks to address that gap by evaluating the use and reported psychometric properties of the ASEBA forms in SSA.

The majority of behavioural screening tools used in SSA are developed in North America or Europe and are generally well established in these regions (Fernald et al., 2009; Sharp et al., 2014; Sweetland et al., 2014). Using such tools in SSA is often more efficient and feasible compared to developing new tools locally, and also enables comparison of findings cross-culturally (Van Widenfelt et al., 2005). However, tools developed in the global north are generally designed for direct application to English-speaking individuals from Western, urbanised, populations (Nezafat Maldonado et al., 2019). It follows that using a tool with individuals who are not English-speaking, or who are from cultures that differ substantially from that of the original target population, may present issues for both administration and interpretation (Sweetland et al., 2014).

There are several challenges associated with translating a tool from English into another language (De Kock et al., 2013; Fernald et al., 2009). For example, an English word or phrase may not have a linguistic equivalent in the target language, or, if an equivalent word or phrase does exist, it may not form part of the vernacular of the target population. A direct translation may also have a slightly different or ambiguous meaning in the target language (Van Widenfelt et al., 2005). Many African languages do not have established terms to describe specific mental illnesses, emotions, or personality traits (Atilola, 2015; Van Eeden & Mantsha, 2007). Poor translations of items that measure psychological constructs may therefore introduce bias, which may, in turn, compromise the validity and reliability of the scores derived from the tool.

It is also important to consider whether constructs being measured by a tool are relevant and understood in the same way in different cultures (i.e., construct equivalence; Van De Vijver & Leung, 2011). This is applicable even when a tool is administered in its original English. A South African study conducted a pilot test to evaluate the cultural appropriateness of items on the Child Behaviour Checklist (CBCL) in English and in two other South African languages (LeCroix et al., 2020; Palin et al., 2009). Feedback from participants led to the removal of the item “sets fires”, intended to measure rule-breaking behaviour. In this context, it is likely that setting fires (e.g., for cooking) is commonplace amongst children and adolescents as part of daily life, and so participants interpreted the item in this way, instead of the intended interpretation (i.e., setting a fire with intent to cause harm or damage). Hence, establishing linguistic and construct equivalence prior to using a tool outside of its original context is critical to ensure that the tool is measuring what it intends to measure.

Measurement, or psychometric, properties (e.g., validity and reliability) are not properties of the tool itself, but are characteristics of the data derived from the tool in a specific context (Zumbo & Chan, 2014). Most applied research studies conduct rudimentary psychometric evaluations of scores obtained from psychological tools (Dima, 2018; Flake et al., 2017; Vacha-Haase & Thompson, 2011). The result is the use of tools, including behavioural screening tools, without sufficient evidence to support the validity and reliability of the scores derived from the tools in a given context. Scores generated from such a tool may not accurately reflect the ‘true scores’ of the respondents (De Kock et al., 2013). This may, in turn, increase the risk of misdiagnosis, which has implications for referral and the provision of appropriate interventions. Hence, until psychometric equivalence of a behavioural screening tool is established, results should be interpreted with caution.

The ASEBA forms, developed in the United States, are currently used as screening tools for clinical and research purposes in SSA. The ASEBA forms are designed to quickly and effectively measure maladaptive behaviours in children and adolescents. One major advantage of the ASEBA forms is that data can be obtained from multiple informants (i.e., caregiver, teacher, and self-report), allowing for a comprehensive overview of the child’s behaviour in different contexts. The most recent version of the Preschool Forms include the parent-report Child Behaviour Checklist for ages 1.5-5 (CBCL/1.5-5) and the Caregiver-Teacher Report Form for Ages 1.5-5 (C-TRF; Achenbach & Rescorla, 2000). The School-Age Forms include the parent-report Child Behaviour Checklist for Ages 6-18 (CBCL/6-18), the Teacher’s Report Form (TRF), and the Youth Self Report (YSR; Achenbach & Rescorla, 2001). In this review, we refer to the parent report forms collectively as the ‘CBCL’, but when referring to a specific age form, we use the corresponding ASEBA abbreviation (e.g., CBCL/1.5-5 or CBCL/6-18).

The forms are presented as lists of items describing a range of behaviours, (e.g., “avoids looking others in the eye”, “has trouble getting to sleep”). Respondents indicate their agreement with the items by selecting either “not true” (scored as a 0), “somewhat or sometimes true” (scored as a 1) or “very true/often true” (scored as a 2). These scores are summed to provide a Total Problems score, where higher scores indicate the presence of more problem behaviours. Items are grouped into syndrome scales, which are further grouped into two broad band scales (Internalising Problems and Externalising Problems, see Figures 1 and 2). Items are also grouped into DSM-oriented scales (see Figures 1 and 2), aligned with diagnostic criteria for a number of disorders specified in the fifth edition of the Diagnostic and Statistical Manual for Mental Disorders (DSM-5; American Psychiatric Association, 2013).

**Figure 1.**
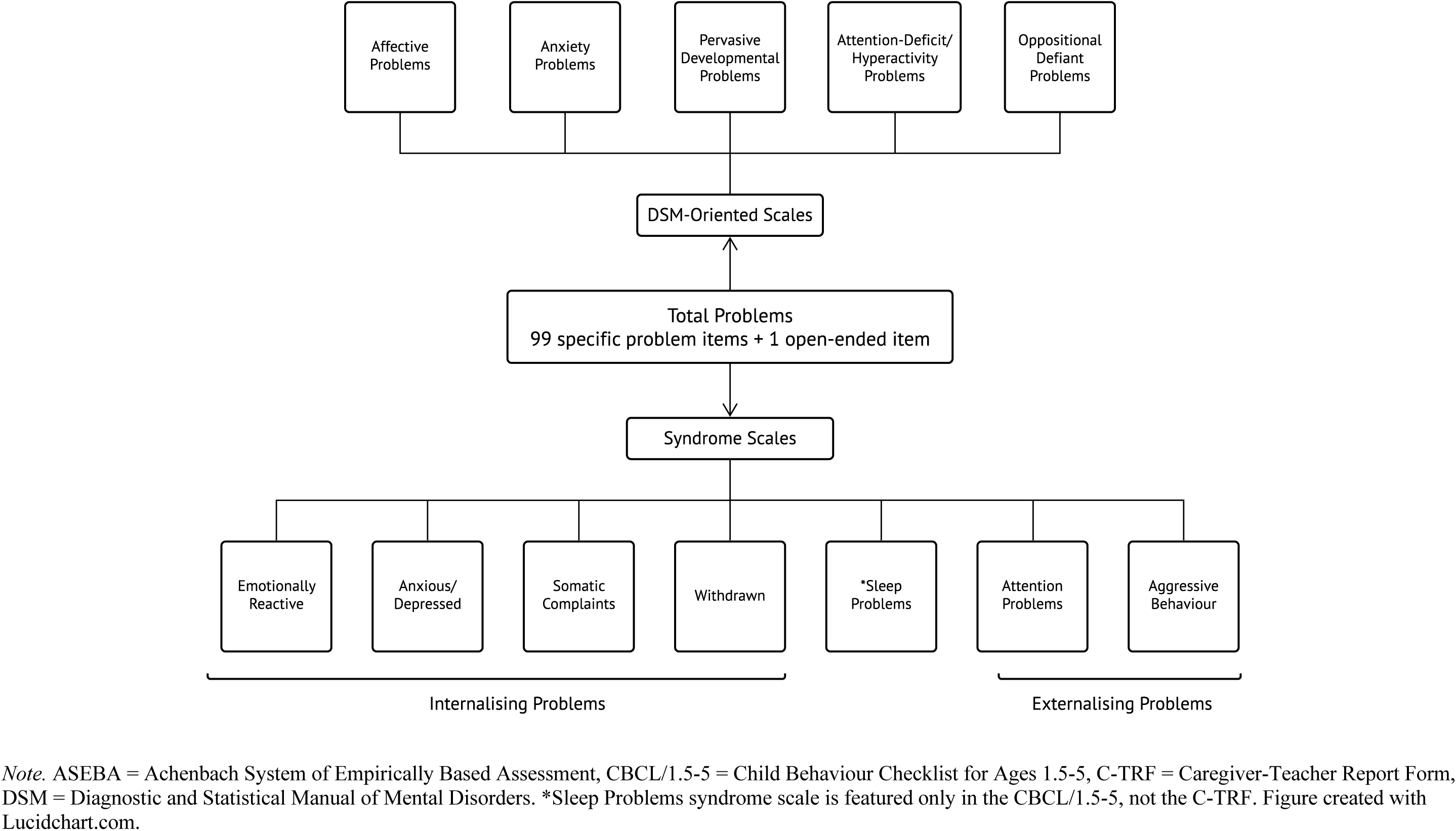
Structure of the ASEBA Preschool Forms (CBCL/1.5-5 and C-TRF): Syndrome Scales and DSM-Oriented Scales

**Figure 2.**
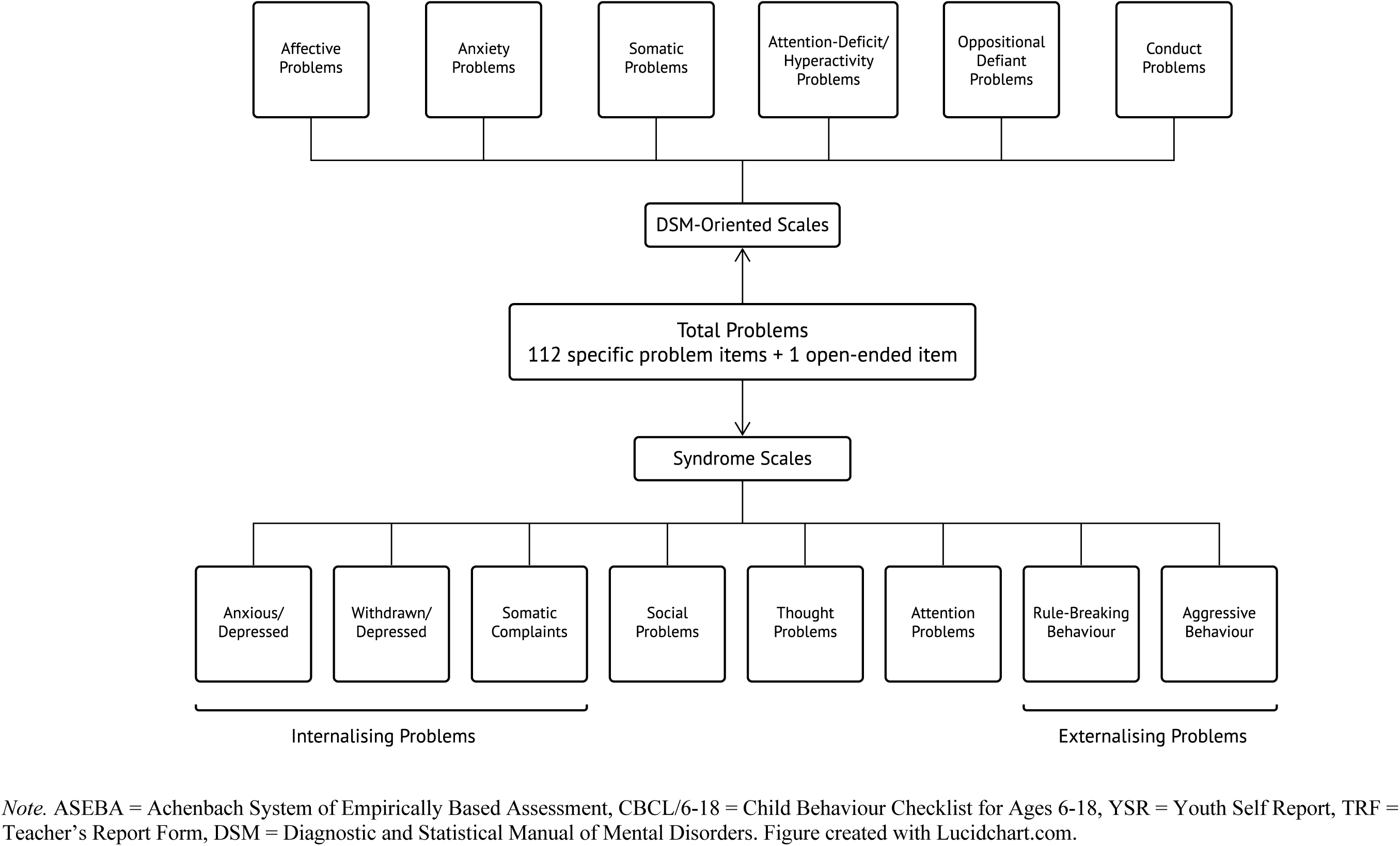
Structure of the ASEBA School-Age Forms (CBCL/6-18, YSR, and TRF): Syndrome Scales and DSM-Oriented Scales

It is not clear where or how the ASEBA forms are used in SSA, or to what extent the scores from the ASEBA forms have been evaluated for their validity, reliability, and cultural appropriateness for the diverse populations and contexts in this region. This study had four primary objectives, namely (i) to collate all studies that used the ASEBA forms with sub-Saharan African (SSAn) participants, (ii) to describe the use of the ASEBA forms across SSA (including the use of translations), (iii) to evaluate the reported psychometric properties of the scores of different forms and subscales, and finally, (iv) to make recommendations regarding the use of the ASEBA forms in SSA based on available evidence.

## Methods

We searched PubMed, EBSCO (APA PsycInfo, APA PsycArticles, ERIC, Academic Search Premier, Health Source: Nursing/Academic Edition, Africa-Wide Information, CINAHL), Scopus, and Google Scholar databases. In addition, we searched ProQuest and the University of Cape Town’s (UCT) library database for relevant dissertations/theses, book chapters, and conference abstracts. A detailed overview of the search strategy is presented in Table S1 in the digital supplement.

### Search Strategy

For all database searches, the ASEBA terms “child behaviour checklist” OR “child behavior checklist” OR CBCL OR “Youth Self-Report” OR “Teacher’s Report Form” were added as the first line of the search. Although most publications used the American spelling (“behavior”), the British spelling was included as an option so as not to exclude articles from journals that use British spelling. A preliminary search including only the ASEBA terms revealed a number of journal articles that referred to the CBCL as the “Children’s Behaviour Checklist”, “Child Behaviour Check List”, or “CBC” to refer to the same tool. However, there was no substantial difference in the number of search results when these variants were included in the final search terms. We conducted another trial search without inverted commas around “Child Behaviour Checklist” to check if authors were using the tool name more loosely. However, many more results showed up and most of them were not meaningful (i.e., included other tools with the word ‘checklist’ in their names). We did not include the abbreviations for the Youth Self-Report (YSR) or the Teacher’s Report Form (TRF). Including “YSR” did not substantially affect the number of results and including “TRF” generated too many irrelevant results.

The SSAn search terms were adapted from Pienaar and colleagues (2011) and the list of SSAn countries from the United Nation’s Standard Country or Area Codes for Statistical Use (United Nations Statistics Division, 1998). We excluded surrounding islands and territories (e.g., Madagascar, Comoros) from the SSAn search terms, as we were primarily interested in continental SSAn countries. For four countries, we included alternative names (e.g., “Ivory Coast” in addition to “Côte d’Ivoire”). After conducting preliminary searches with the ASEBA and SSA terms described above, we noticed that many results were coming up with African-American samples. Hence, for all searches, we narrowed the search by excluding the following terms: “African-American” OR “African American”.

We did not include any psychometrics-related words or terms (e.g., validity, Cronbach’s alpha) because of (i) the broad range of psychometric-related terms, and (ii) inconsistencies in indexing and reporting of psychometric properties. As it was more likely that a study utilised the CBCL as an outcome measure rather than the tool itself being the subject of the study, no Medical Subject Headings (MeSH terms) were included in the search terms. For the same reason, “all/full text” fields were selected for all lines of the searches, except for one database where it yielded too many results (see Table S1 in the digital supplement). No coverage dates (i.e., year limits) were selected in any database search, nor was the type of publication. All records were saved to a reference library (Endnote X9), after which duplicate records were removed.

### Inclusion Criteria

A study was eligible to be included in the final analysis, if:

i. The study was written in English.
ii. The study reported original findings.
iii. The study sample (or at least a portion of the sample) was from a SSAn country. Immigrants and refugees currently living outside SSA were eligible if the study reported specific data for the SSAn participants. For immigrants/refugees, either the child or at least one parent/caregiver had to have been born in a SSAn country.
iv. The study used an ASEBA form (any form or any version) in its standard format (i.e., as per the official instructions in the ASEBA manual), and reported the data derived from the tool. Minor adaptations to the tool (e.g., excluding items due to cultural inappropriateness etc.) were acceptable, as long as the modifications were clearly specified and justified, and the tool was still recognisable as an ASEBA form.
v. The study reported psychometric properties (e.g., validity, reliability) of the ASEBA form for the study sample. Inherent in this criterion is the exclusion of case studies.

### Screening and Review Process

Two of the authors (MRZ and CF) independently screened and reviewed all records for eligibility. As the ASEBA forms were generally not the subject of the study (and therefore did not appear in the title or abstract), the full text of each article was scanned or read at each stage of the review process until relevance or eligibility (or lack thereof) became clear. At each stage, the reviewers identified and discussed any discrepancies until a consensus was reached. In the event of an impasse, the reviewers presented the article in question to the fourth author (KAD), who made the final decision.

The review comprised three distinct stages:

i. A brief screening of all full-text studies to check for relevance. Did the study include a SSAn sample and use an ASEBA form?
ii. A more thorough screening of the relevant studies to check for eligibility. Did the study describe the SSAn sample (or sub-sample) and were there specific data for those participants? Was the ASEBA form used in the standard way? We excluded studies if the description of the sample or the country of origin was vague or if there were no specific data pertaining to the SSAn participants. We also excluded studies that used an ASEBA form in a non-standard way, as we wanted to reasonably compare the psychometric properties across studies. At this stage, the reviewers scanned the reference lists of relevant articles to look for other literature that did not appear in the original search results.
iii. A review of the studies identified at the second stage to determine if the study reported psychometric properties of the tool. Any psychometric analyses were acceptable, so long as the statistics were for the study sample (i.e., not from another study or the tool manual). We included studies that met these criteria in the final analysis.

We extracted and summarised key information from the included studies, such as details related to the sample, the country of origin, the ASEBA versions administered, the language(s) of administration, any translation or adaptation processes, and the psychometric analyses conducted. If any details were missing from an article or were unclear, we contacted the corresponding author for clarification or, if applicable, referred to other articles related to the same umbrella study. In the event that the corresponding author did not respond after two attempts to contact them, we noted the uncertainty in our records.

We then evaluated the psychometric properties of the ASEBA forms with reference to COSMIN (COnsensus-based Standards for the selection of health Measurement INstruments) criteria for good measurement properties (Prinsen et al., 2018). COSMIN describes three phases of psychometric evaluation. The first phase involves investigating a tool’s content validity, that is, the extent to which the content of the tool adequately reflects the construct being measured. Specifically, COSMIN recommends that tools be relevant, comprehensive, and comprehensible with respect to the construct of interest and the target population (Terwee, Prinsen, Chiarotto, Westerman, et al., 2018). The second phase concerns evaluating the internal structure of the tool, including structural validity, internal consistency, and cross-cultural validity (measurement invariance). The third phase involves evaluating the remaining measurement properties, including reliability, measurement error, criterion validity, and hypothesis testing for construct validity, including concurrent, convergent, divergent, and known-groups validity (Mokkink, Prinsen, et al., 2018). Each measurement property is rated on a three-point scale of ‘sufficient’, ‘indeterminate’, or ‘insufficient’.

We also assessed the methodological quality of the studies using the COSMIN Risk of Bias checklist (Mokkink, de Vet, et al., 2018). COSMIN utilises a four-point rating system to grade each measurement property in a study as ‘very good’, ‘adequate’, ‘doubtful’ or ‘inadequate’. The overall rating of the quality of each measurement property is determined by taking the lowest rating of any standards corresponding to that property.

## Results

### Search Results

A flow diagram adapted from the Preferred Reporting Items for Systematic Reviews and Meta-Analyses (PRISMA) Statement (Moher et al., 2010), details the number of studies included and excluded at each stage of the review process (Figure 3).

**Figure 3.**
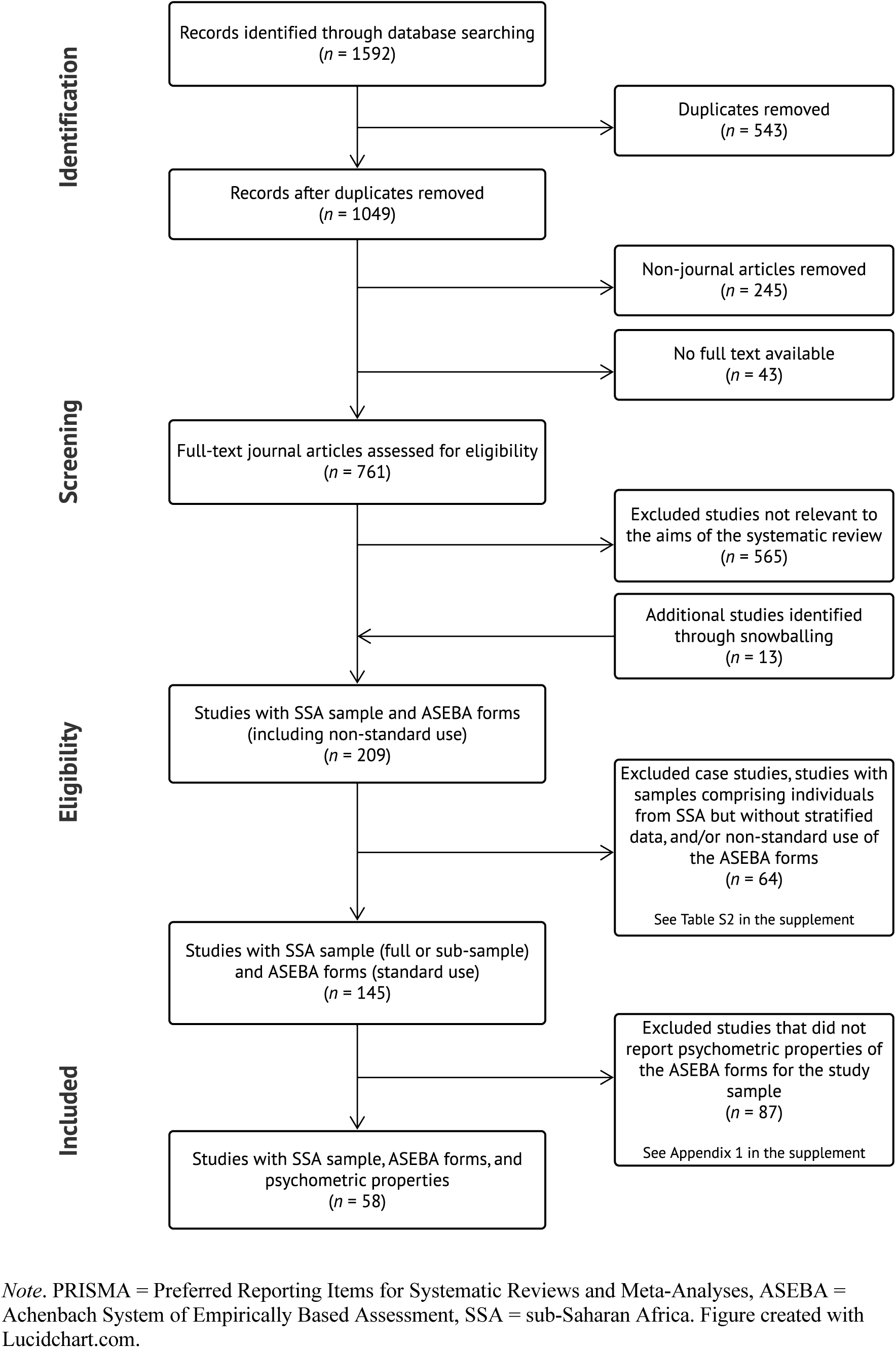
PRISMA Diagram Adapted from Moher et al. (2010) Outlining the Number of Records Included and Excluded at Each Phase of the Systematic Review

During the search for manuscripts, it became apparent that full-text versions of many “grey” references, including conference abstracts, poster presentations, dissertations/theses, and other unpublished work, were difficult to access. We decided to randomly sample 20 records from each category of grey literature to check for relevance. The aim of this test was to determine if it was worth pursuing the full-text search for these records. Out of the 20 conference abstracts/presentations sampled, only one was relevant to the aims of the systematic review. The dissertation category was more promising in terms of relevance. However, we could not access full texts for over 60% of the records in these categories. We experienced similar difficulties when trying to access full-text versions of book chapters (digital and/or hard copies). Considering all these factors, we decided to exclude 245 records of “grey” literature and book chapters, and to focus exclusively on published journal articles. These included articles from peer reviewed monographs or reports from reputable institutions with original data, as these papers were easily accessible and contained relevant and useful data.

We could not access the full texts of 43 out of the 804 journal articles (5%); these records were therefore excluded. The remaining 761 full-text articles were screened for relevance. Five hundred and sixty-five records (74%) were deemed irrelevant to the aims and scope of the systematic review. During the screening process, the reviewers discovered 13 possibly relevant journal articles that were not included in the search results. This resulted in a total of 209 journal articles with (possible) SSAn participants who completed the ASEBA forms. Of the 209 studies, 64 (31%) were excluded for reasons related to the sample description and use of the ASEBA forms. Specifically, 20 studies described the African sample vaguely (e.g., “participants from Africa”, with no specific reference to country of origin), and a further 20 studies included SSAn participants in their samples but did not report stratified ASEBA data for those participants. Furthermore, 19 studies used or scored items in non-standardised ways. Examples of non-standardised use included administering a small selection of items independently (i.e., not as part of an established ASEBA subscale), using an incomplete subscale without justification, using a few items as part of a new measure, and altering the standard response format (e.g., from a three-point scale to a two-point scale). We did not have access to ASEBA forms published before 1991, so we could not ascertain whether the three studies that used these older versions administered full or partial subscales. Two studies (reporting data from the same sample) used a modified version of the YSR whose items bore little resemblance to those of the original YSR. Two studies administered the ASEBA forms as part of the study but did not report the actual data. Finally, we excluded two case studies. A brief description of these excluded articles can be found in Table S2 in the digital supplement.

After these exclusions, we were left with 145 studies with specific data for SSAn participants that used the ASEBA forms in the standardised way. A few different articles stemmed from the same ‘umbrella’ study, as indicated by identical or overlapping samples. In addition, a few multi-nation studies utilised the same data set for secondary data analysis. Of the 145 studies, only 58 (40%) reported the psychometric properties of the ASEBA forms for the study sample and these studies were included in the final analysis. The digital supplement presents a list of the studies without reported psychometric properties (*n* = 87; Appendix S1).

### Study Characteristics

#### Region

Figure 4 illustrates the number of studies from the different SSAn countries. The map presents data from all 145 studies that met the first two inclusion criteria (i.e., SSAn participants and standardised use of the ASEBA forms), regardless of whether psychometric properties were reported. We displayed data from all 145 studies to visualise patterns of ASEBA usage across SSA. Six studies from outside of SSA comprised immigrant participants who originated from one of the following fourteen countries: Angola, Eritrea, Ethiopia, Gambia, Ghana, Guinea, Guinea-Bissau, Kenya, Mali, Niger, Nigeria, Sierra-Leone, Senegal, and Somalia.

**Figure 4.**
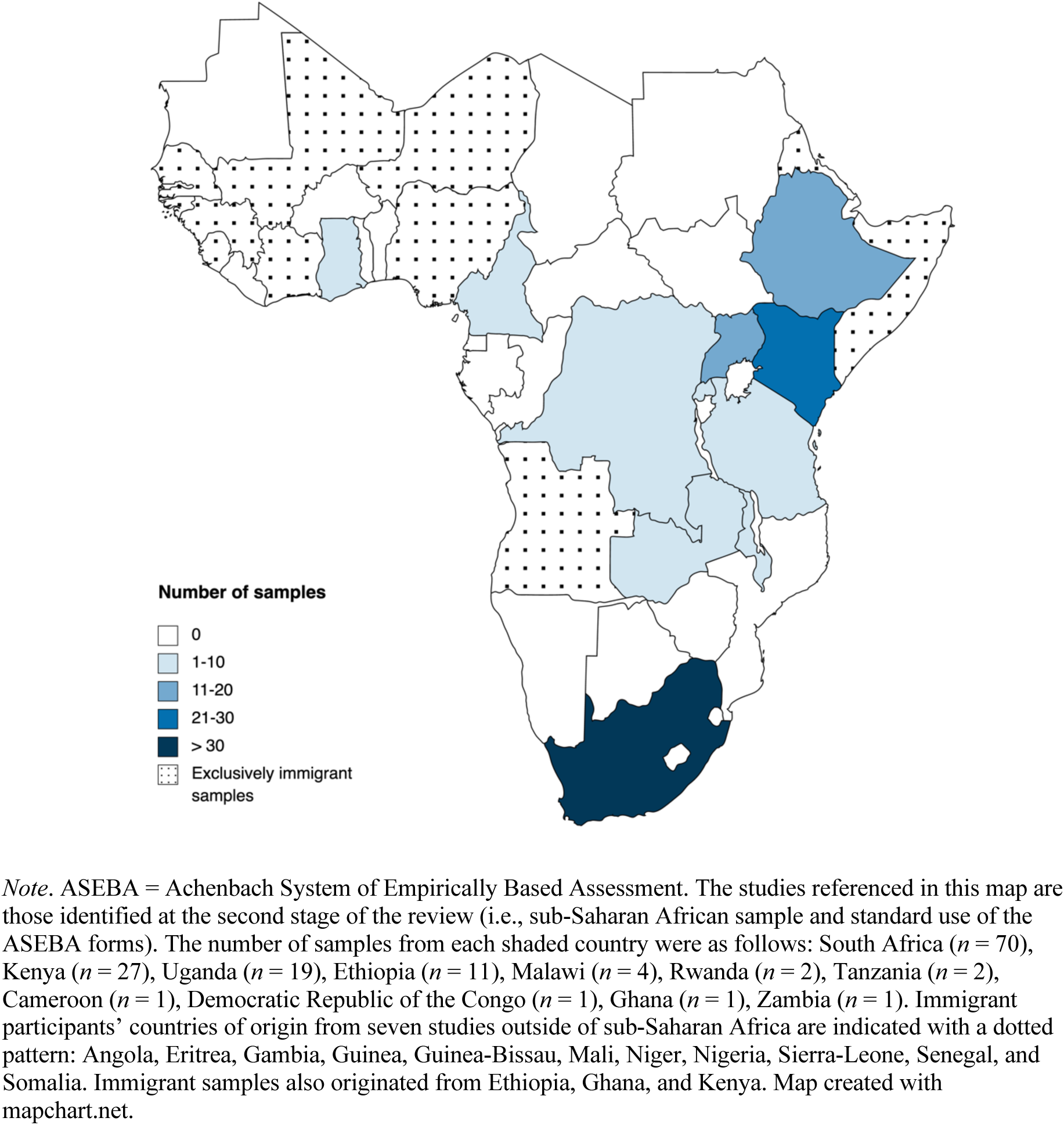
Map of Sub-Saharan Africa Displaying Number of ASEBA Studies Originating from Each Country (N = 145)

An overview of the map shows the predominance of studies with samples from Southern African (*n* = 70, 48%) and East African (*n* = 66, 45%) countries. In Southern Africa, all samples originated from South Africa, while East African samples were distributed across a number of countries, including Kenya (*n* = 27, 41%), Uganda (*n* = 19, 29%), Ethiopia (*n* = 11, 17%), Malawi (*n =* 4, 6%), Rwanda (*n* = 2, 3%), Zambia (*n* = 1, 2%), and Tanzania (*n* = 1, 2%). In contrast, there were only two studies from Central Africa, which came from the Democratic Republic of the Congo (DRC) and Cameroon respectively, and only one study from West Africa, which came from Ghana. Many participants from immigrant samples originally came from countries in West Africa.

The distribution of studies across SSA was similar after narrowing the studies down to those that reported psychometric properties of the CBCL (*N* = 58). The breakdown of studies by region was as follows: Southern Africa (*n* = 29, 50%), East Africa (*n* = 25, 43%), Central Africa (*n* = 1, 2%) and immigrant samples (*n* = 3, 5%) from East and West Africa, living in Sweden (*n* = 1) and the USA (*n* = 2).

Relevant information about the studies including the reported psychometric properties (*n* = 58) is presented in Table 1.

**Table 1.** Description of Studies that Administered the ASEBA Forms to a Sub-Saharan African Sample and Reported their Psychometric Properties (N = 58)

| Author(s) | Sample description | Version | Language(s) of administration | Translation/adaptation process | Purpose of the tool in the context of study aims | Psychometric properties |
| --- | --- | --- | --- | --- | --- | --- |
| Southern Africa (*n* = 29) |  |  |  |  |  |  |
| South Africa (*n* = 29) |  |  |  |  |  |  |
| Allen et al. (2013) | HIV negative children (aged 6-10 years) with HIV positive mothers, living in a township community in Tshwane<br>*N* = 361 | CBCL/4-18 (1991) | Sepedi, isiZulu, Setswana, Sesotho, and English | • Translation<br>• Back-translation<br>• Reviewed by cultural advisors (community representatives)<br>• Pilot test (*N* = 20) | Investigated relationships between maternal psychological functioning, parenting, and child behaviour in an HIV/AIDS context. | <u>IC (Cronbach's $\alpha$)</u><br>Internalising Problems [$\alpha$ = 0.852]<br>Externalising Problems [$\alpha$ = 0.915] |
| aBarber et al. (2005)<br>*Linked to Bradford et al. (2003)* | Xhosa- (*n* = 600), Afrikaans- (*n* = 600), and English-speaking (*n* = 600) adolescents (aged 13-17 years) living in Cape Town | YSR (1987) | Xhosa, Afrikaans, and English | • Translation<br>• Back-translation<br>• Reviewed by community collaborators | Analysed links between parenting and adolescent functioning across countries/ethnic groups. | <u>IC (Cronbach's $\alpha$)</u><br>*Delinquent Behaviour (8 items)*<br>Xhosa [$\alpha$ = 0.75]<br>Afrikaans [$\alpha$ = 0.79]<br>English [$\alpha$ = 0.75] |
| Boyes et al. (2012); Boyes and Cluver (2013) | Children and adolescents, (aged 10-19 years) living in peri-urban communities in Cape Town<br>*N* = 1025 | YSR (1991) | Xhosa | • Translation<br>• Back-translation<br>• Reviewed by 5 health and social workers | Determined the psychometric properties of the Revised Children's Manifest Anxiety Scale and the Child PTSD Checklist. | <u>IC (Cronbach's $\alpha$)</u><br>Somatic Complaints (9 items) [$\alpha$ = 0.66]<br>Delinquent Behaviour [$\alpha$ = 0.61] |
| aBradford et al. (2003) | Xhosa- (*n* = 635), Afrikaans- (*n* = 520), and English-speaking (*n* = 579) adolescents (aged 14-17 years) living in Cape Town | YSR (1987) | Xhosa, Afrikaans, and English | • Translation<br>• Back-translation<br>• Reviewed by community collaborators | Investigated the relationship between interparental conflict and child outcomes in nine countries. | <u>IC (Cronbach's $\alpha$)</u><br>*Delinquent Behaviour (6 items)*<br>Xhosa [$\alpha$ = 0.75]<br>Afrikaans [$\alpha$ = 0.72]<br>English [$\alpha$ = 0.70] |
| #Cluver, Lachman, et al. (2017) | Adolescents (aged 10-17 years) living in two poor rural communities in the Eastern Cape province<br>*N* = 30 | CBCL and YSR (1991) | Xhosa | • Translation<br>• Back-translation | Evaluated the effectiveness of a parenting programme. Problem behaviour was measured pre- and post-intervention. | <u>IC (Cronbach's $\alpha$)</u><br>*Rule-Breaking Behaviour (17 items)*<br>CBCL [$\alpha$ = 0.82]<br>YSR [$\alpha$ = 0.62]<br>*Aggressive Behaviour (18 items)*<br>CBCL [$\alpha$ = 0.85]<br>YSR [$\alpha$ = 0.54] |
| Cluver, Meinck, Yakubovich, et al. (2016) | Adolescents (aged 10-17 years) living in poorly resourced rural and peri-urban communities.<br>*N* = 115 (dyads) | CBCL/4-18 and YSR (1991) | Xhosa | • Translation<br>• Back-translation | A pre-post trial testing the effectiveness of a parenting support programme to reduce child abuse. | <u>IC (Cronbach's $\alpha$)</u><br>*Externalising Problems (35 items)*<br>CBCL [$\alpha$ = 0.88]<br>YSR [$\alpha$ = 0.71] |
| Collishaw et al. (2016) | Children and adolescents (aged 10-19) living in urban settlements in Cape Town<br>Time 1: *N* = 1025<br>Time 2: *N* = 716 | YSR (1991) | Xhosa | • Translation<br>• Back-translation<br>• Pilot test | Identified predictors of mental health resilience in children parentally bereaved by AIDS. | <u>IC (Cronbach's $\alpha$)</u><br>*Delinquency (11 items)*<br>Time 1 [$\alpha$ = 0.61]<br>Time 2 [$\alpha$ = 0.64] |
| Cortina et al. (2013) | Primary school-going children (aged 10-12 years) from a poor rural area in Mpumalanga province<br>*N* = 1025 | YSR (1991) | Shangaan | • Translation<br>• Back-translation<br>• Pilot test (*N* = 200) | Estimated the prevalence of psychological problems amongst children in a poor rural school setting. | <u>IC (Cronbach's $\alpha$)</u><br>Anxious/Depressed (13 items) [$\alpha$ = 0.63] |
| du Plessis et al. (2015) | Primary school children (aged 12-15 years) living in low-income communities in Cape Town<br>*N* = 616 | YSR (1983) | English and Afrikaans | • Translation<br>• Back-translation<br>• Pilot test (*N* = 72) | Examined associations between exposure to violence and internalising and externalising behaviours. | <u>IC (Cronbach's $\alpha$)</u><br>*Delinquency (12 items)*<br>Full sample [$\alpha$ = 0.70]<br>English version [$\alpha$ = 0.67]<br>Afrikaans translated version [$\alpha$ = 0.75] |
| Gwandure (2007) | School-going children (aged 14-19 years) providing home-based AIDS care for a parent/caregiver ($n = 30$) and a control group ($n = 30$) | CBCL/4-18 (1991) | NR | NR | Investigating the impact of providing home-based AIDS care on children's psychological and cognitive functioning. | <u>IC (Cronbach's $\alpha$)</u><br>Total Problems [$\alpha = 0.87$] |
| LeCroix et al. (2020)<br>*Linked to Palin et al. (2009)* | Children (aged 11-16 years) with a mother living with HIV<br>$N = 104$ | CBCL (1991) | English, Afrikaans, and Sotho | • Translation<br>• Back-translation<br>• Reviewed by community collaborators<br>• Pilot test and focus groups | Investigated the role of mother-child relationships in the context of maternal HIV and child outcomes. | <u>CFA</u><br>*Internalising Problems*<br>The Somatic Complaints subscale was removed as the items did not load adequately. An additional 5 items were eliminated due to translation errors during the back-translation process. Of the remaining 18 items, 13 items loaded at 0.40 and above.<br>*Externalising Problems*<br>One item (i.e., sets fires) was excluded given feedback received from focus groups and piloting. Of the remaining 32 items, 23 items loaded at 0.40 and above.<br><u>IC (Cronbach's $\alpha$)</u><br>Internalising Problems (13 items) [$\alpha = 0.77$]<br>Externalising Problems (23 items) [$\alpha = 0.87$] |
| Malcolm-Smith et al. (2015) | Grade 1 learners (aged 6-8 years) from an English-medium school serving a predominantly lower middle-class community in Cape Town<br>$N = 65$ | CBCL (2001) | English | NA | Investigated associations between empathy and aggression in young school-going children. | <u>IC (Cronbach's $\alpha$)</u><br>Externalising Problems [$\alpha = 0.87$] |
| #Meinck et al. (2018)<br>*Linked to Shenderovich et al. (2020)* | Adolescents (aged 10-18 years), living in the Eastern Cape Province<br>*N = 552* | YSR (2001) | Xhosa | • Translation<br>• Back-translation<br>*Extracted from Chuver, Meinck, Shenderovich, et al. (2016)* | Investigated the psychometric properties of a child abuse self-report screening tool. | <u>IC (Cronbach's $\alpha$)</u><br>Externalising Problems [$\alpha = 0.85$] |
| #Meinck et al. (2017) | Adolescents (aged 10-17 years) from rural and urban districts in KwaZulu-Natal province<br>*N = 2477* | YSR (2001) | IsiZulu | • Translation<br>• Back-translation | Investigated the relationship between family-related factors (e.g., deprivation, parenting) and adolescent health outcomes. | <u>IC (Cronbach's $\alpha$)</u><br>Delinquency [$\alpha = 0.64$] |
| #Meinck et al. (2019) | Adolescent girls (aged 10 to 17 years) from urban and rural areas in two South African provinces<br>*N = 3516* | YSR (2001) | Zulu, Xhosa, Sotho, Sepedi, Swati and Tsonga | • Translation<br>• Back-translation | Investigated the moderating effects of free schooling in the relationship between adverse childhood experiences and HIV risk behaviours. | <u>IC (Cronbach's $\alpha$)</u><br>Delinquency [$\alpha = 0.58$] |
| Palin et al. (2009) | Children (aged 6-11 years) whose primary caregivers were woman living with HIV<br>*N = 103* | CBCL/4-18 (1991) | Sotho, English, and Afrikaans | • Translation<br>• Back-translation<br>• Pilot test and focus groups | Studied the relationship between disclosure of maternal HIV infection and child behaviour. | <u>CFA</u><br>Somatic complaints items were eliminated for analyses, as factor loadings for these items were low ($<0.40$). An additional five items were eliminated due to translation errors. One item ("Sets fires") was excluded based on feedback from piloting and focus group.<br><u>IC (Cronbach's $\alpha$)</u><br>Internalising Problems (13 items) [$\alpha = 0.78$]<br>Externalising Problems (23 items) [$\alpha = 0.88$] |
| Peltzer and Pengpid (2013) | Children (aged 1.5-5 years) with a mother who had obtained a protection order against a male partner in Vhembe district, Limpopo province<br>*N* = 86 | CBCL/1.5-5 (2000) | Tsonga and Venda | • Translation<br>• Back-translation | Examined associations between intimate partner violence and behavioural problems among pre-school children. | <u>IC (Cronbach's $\alpha$)</u><br>Internalising Problems [$\alpha$ = 0.92]<br>Externalising Problems [$\alpha$ = 0.94]<br>Total Problems [$\alpha$ = 0.96] |
| #Rawatlal et al. (2015) | School-going adolescents (aged 9-18 years) in KwaZulu Natal province<br>*N* = 206 | CBCL/4-18 (1991) | English | NA | Examined the relationship between adolescent-parent attachment, perceived support from parents, and internalising symptoms. | <u>IC (Cronbach's $\alpha$)</u><br>Depression/Anxiety (14 items) [$\alpha$ = 0.81] |
| #Rochat, Houle, et al. (2017) | HIV-exposed and unexposed children (aged 7-11 years) from Hlabisa sub-district in KwaZulu-Natal province<br>*N* = 1536 | CBCL/6-18 (2001) | Zulu | • Translation<br>• Back-translation<br>• Obtained translation license from ASEBA and translation was approved by test developer | Examined associations between early life factors, a breastfeeding intervention, and later child emotional and behavioural problems. | <u>IC (Cronbach's $\alpha$)</u><br>Total Problems [$\alpha$ = 0.94] |
| #Rochat et al. (2015);<br>#Rochat, Mitchell, et al. (2017) | HIV-uninfected children (aged 6-10 years) of HIV-infected women from the Hlabisa sub-district in KwaZulu Natal province<br>*N* = 281 | CBCL/6-18 (2001) | Zulu | • Translation<br>• Back-translation<br>• Obtained translation license from ASEBA and translation was approved by test developer | Examined child psychological outcomes of maternal HIV disclosure intervention and communication about HIV and death in a rural South African setting. | <u>IC (Cronbach's $\alpha$)</u><br>*Total Problems*<br>Pre-intervention [$\alpha$ = 0.94]<br>Post-intervention [$\alpha$ = 0.92] |
| #Rochat et al. (2016) | HIV-negative children (aged 7-11 years) born to HIV-positive and HIV-negative mothers<br>*N* = 906 | CBCL/6-18 (2001) | Zulu | • Translation<br>• Back-translation<br>• Obtained translation license from ASEBA and translation was approved by test developer | Investigated the relationship between exclusive breastfeeding and child behavioural problems, in HIV-uninfected (exposed and unexposed) children. | <u>IC (Cronbach's $\alpha$)</u><br>Total Problems [$\alpha$ = 0.94] |
| Shenderovich et al. (2020) | Adolescents (aged 10-18 years) living in the Eastern Cape<br>*N* = 552 | CBCL (2000) | Xhosa | • Translation<br>• Back-translation<br>*Extracted from Chuver, Meinck, Shenderovich, et al. (2016)* | Studied moderators of outcomes of a parenting intervention programme. | <u>IC (Cronbach's $\alpha$)</u><br>Externalising Problems (35 items) [$\alpha$ = 0.90] |
| #Sipsma et al. (2013) | Young children (aged 6-10) with HIV-positive and HIV-negative mothers, living in Tshwane<br>*N* = 509 | CBCL/6-18 (2001) | Likely Sepedi, Setswana, Sesotho, isiZulu and English | • Translation<br>• Back-translation<br>• Reviewed by community advisors<br>• Pilot test (*N* = 20) | Compared behaviour and psychological functioning of children with HIV-positive and HIV-negative mothers. | <u>IC (Cronbach's $\alpha$)</u><br>Internalising Problems ($\alpha$ = 0.85)<br>Externalising Problems ($\alpha$ = 0.92) |
| #Swain et al. (2017) | School-going adolescents (aged 9-18 years) living in a low socioeconomic community in KwaZulu Natal province<br>*N* = 324 | CBCL/4-18 (1991) | English | NA | Estimated the prevalence of posttraumatic stress in adolescents living in low-socioeconomic communities. | <u>IC (Cronbach's $\alpha$)</u><br>Anxiety/Depression [$\alpha$ = 0.81]<br>Somatic Complaints [$\alpha$ = 0.77] |
| #aThornton et al. (2019) | Children/adolescents (aged 11-18 years, 67% orphaned)<br>*N* = 175 | YSR (2001) | English | NA | Identified clinical risk factors amongst young people at risk of suicide, living in South Africa and Guyana. | <u>IC (Cronbach's $\alpha$)</u><br>Attention Problems (10 items) [$\alpha$ = 0.58];<br>Anxious/Depressed (8 items) [$\alpha$ = 0.60];<br>Withdrawn/Depressed (14 items) [$\alpha$ = 0.79];<br>Somatic Complaints (11 items) [$\alpha$ = 0.77];<br>Thought Problems (10 items) [$\alpha$ = 0.75];<br>Social Problems (11 items) [$\alpha$ = 0.73] |
| #van Westrhenen et al. (2019) | Children (aged 7-13 years) who experienced a traumatic event or abuse, living in and around Johannesburg<br>*N* = 125 | CBCL/4-18 (1991) | English | NA | Evaluated a creative arts psychotherapy group programme with children who experienced a trauma. | <u>IC (Cronbach's $\alpha$)</u><br><u>Total Problems</u><br>Pre-intervention (*n* = 125) [$\alpha$ = 0.96]<br>Post-intervention (*n* = 37) [$\alpha$ = 0.96] |
| #Visser et al. (2018) | HIV-infected and HIV-uninfected children (aged 6-12 years) living in Tshwane<br>*N* = 167 | CBCL/6-18 (2001) | Sepedi, Setswana, Sesotho and isiZulu | • Translation<br>• Back-translation<br>• Reviewed by community advisors<br>• Pilot test (*N* = 20) | Compared psychological problems of HIV-infected and HIV-uninfected children. | <u>IC (Cronbach's $\alpha$)</u><br>Affective Problems (13 items) [$\alpha$ = 0.73]; Anxiety Problems (6 items) [$\alpha$ = 0.48]; Somatic Problems (7 items) [$\alpha$ = 0.57]; ADHD (7 items) [$\alpha$ = 0.74]; Oppositional Behaviour (5 items) [$\alpha$ = 0.69]; Conduct Problems (17 items) [$\alpha$ = 0.86]; Total DSM-oriented scales (55 items) [$\alpha$ = 0.91] |
| East Africa (*n* = 25) |  |  |  |  |  |  |
| Kenya (*n* = 8) |  |  |  |  |  |  |
| <sup>a</sup>Alampay et al. (2017) | Luo children (aged 7-10 years)<br>*N* = 95 | CBCL/4-18 (1991) | Luo | • Translation<br>• Back-translation | Investigated whether justice and severity moderated the effects of corporal punishment on child internalising and externalising behaviours. | <u>IC (Cronbach's $\alpha$)</u><br>*Internalising (31 items)*<br>Mother-report (*n* = 94) [$\alpha$ = 0.794]<br>Father report (*n* = 90) [$\alpha$ = 0.759]<br>*Externalising (33 items)*<br>Mother-report [$\alpha$ = 0.807]<br>Father-report [$\alpha$ = 0.807] |
| <sup>a</sup>Gershoff et al. (2010) | Children (aged 6-10 years) from the Rachuonyo District of Nyanza province<br>*N* = 17 | CBCL and YSR (1991) | Dholuo | • Translation<br>• Back-translation | Investigated the relationship between parenting discipline practices and child behaviours. | <u>IC (Cronbach's $\alpha$)</u><br>*Aggression:*<br>CBCL (20 items) [$\alpha$ = 0.73]<br>YSR (19 items) [$\alpha$ = 0.81]<br>*Anxiety/Depression:*<br>CBCL (14 items) [$\alpha$ = 0.64]<br>YSR (16 items) [$\alpha$ = 0.81] |
| *Harder et al. (2014) | Children/adolescents (aged 11-18 years) from a poor, informal settlement and surrounding areas in Nairobi<br>*N* = 301 | YSR (2001) | English and Kiswahili | • Translation<br>• Back-translation<br>• Approved by tool developer | Investigated the psychometric properties and factor structure of the YSR in a sample of Kenyan youth. | <u>CFA, IC (Cronbach's $\alpha$)</u><br>*See Table 5.* |
| *Kariuki et al. (2016) | Children (aged 1–6 years) living in Kilifi<br>*N* = 301 | CBCL/1.5-5 (2000) | KiSwahili | • Obtained translation licence from ASEBA<br>• Translation<br>• Back-translation<br>• Translations evaluated by 5 KiSwahili speakers<br>• Focus groups and interviews (*N* = 90)<br>• Pilot test (*N* = 50) | Investigated the psychometric properties of the pre-school version of the CBCL in a Kenyan sample | <u>CFA</u><br>Separate CFAs were conducted for each syndrome scale and for the two broad band scales. RMSEAs for all scales were < 0.08. Mean item factor loadings for syndrome scales ranged from 0.38 (Withdrawn) to 0.53 (Anxious/Depressed).<br><u>IC (Cronbach's $\alpha$)</u><br>Total Problems ($\alpha$ = 0.95)<br>Internalising Problems ($\alpha$ = 0.87)<br>Externalising Problems ($\alpha$ = 0.86)<br>Syndrome scales ($\alpha$ range = 0.53 – 0.86)<br><u>TRR (Pearson's *r*)</u><br>Test-retest reliability of Total Problems (*n* = 38) was significant $r$ = 0.76, $p$ < 0.0001, as was inter-informant agreement between mother and alternative caretaker (*n* = 17; $r$ = 0.89, $p$ < 0.001) and mother and father (*n* = 29; $r$ = 0.81, $p$ < 0.001). |
| Kumar et al. (2018) | Children/adolescents (aged 2-16 years) from two informal communities in Nairobi<br>*N* = 394 | CBCL/1.5-5 (2000) and CBCL/6-18 (2001) | KiSwahili (99%) and English | • Obtained permission from test developer<br>• Translation<br>• Back-translation | Examined the role of maternal adverse childhood experiences on child mental health | <u>IC (Cronbach's $\alpha$)</u><br>Internalising Problems ($\alpha$ = 0.76 – 0.85)<br>Externalising Problems ($\alpha$ = 0.87 – 0.89)<br>*Different alpha statistics were calculated for the different age groups.* |
| *Magai et al. (2018) | Children/adolescents (aged 6-18 years) living in Kenya's Central Province<br>*N* = 1022 | CBCL/4-18 and YSR (1991) | Swahili | • Translated version obtained from ASEBA | Estimated the prevalence of emotional and behavioural problems in Kenyan youth. | <u>CFA, IC (Cronbach's $\alpha$)</u><br>*See Tables 4 and 5.* |
| #Magai et al. (2021) | School-aged survivors of neonatal insults living in Kilifi, Kenya, and a comparison group (aged 6-12 years)<br>*N* = 375 | CBCL/6-18 (2001) | Kiswahili | Translated version obtained from ASEBA | Examined the mental health and quality of life of child survivors of neonatal insults, and a control group. | <u>IC (Cronbach's $\alpha$)</u><br>Alpha coefficients for Internalising Problems, Externalising Problems, and Total Problems ranged from 0.66-0.87 |
| #Skinner et al. (2014) | Luo children (average age 8.46 years, *SD* = 0.64) from Kisumu<br>*N* = 100 | YSR (1991) | Dholuo | Translation<br>Back-translation | Investigated childrearing violence and child adjustment after exposure to post-election violence in Kenya | <u>IC (Cronbach's $\alpha$)</u><br>*Externalising Problems (30 items)*<br>Time 1 [$\alpha$ = 0.72]<br>Time 2 [$\alpha$ = 0.88]<br>Time 3 [$\alpha$ = 0.75] |
| Kenya and Tanzania (*n* = 1) |  |  |  |  |  |  |
| Dorsey et al. (2020) | Children (aged 7-13 years) who had experienced the death of a parent, living in urban and rural sites in Kenya (*n* = 230) and Tanzania (*n* = 230) | CBCL/6-18 (2001) and YSR (1991) | Kiswahili | Translation<br>Back-translation | Tested the effectiveness of a trauma-focused cognitive behavioural therapy for children who experienced the death of a parent. | <u>IC (Cronbach's $\alpha$)</u><br>Alpha coefficients for the CBCL and YSR Internalising Problems and Externalising Problems broadband scales ranged from 0.73-0.89)<br>*The YSR was completed by children aged 11 years or older.* |
| Ethiopia (*n* = 7) |  |  |  |  |  |  |
| <sup>b</sup>Betancourt et al. (2012) | Kunama refugees (aged 11-18 years) participating in an emergency education programme<br>*N* = 153 | YSR and CBCL (1991) | Kunamenga | Translation<br>Back-translation<br>Discussions with community leaders and key informants<br>A few items were removed (e.g., items related to suicide and self-harm) based on community feedback | Assessed adolescent behavioural problems in the context of an education and psychosocial support programme for refugees. | <u>IC (Cronbach's $\alpha$)</u><br>Internalising Problems (29 items) [$\alpha$ = 0.62]<br>Externalising Problems (33 items) [$\alpha$ = 0.77]<br>*Note. It is not clear whether the above statistics correspond to the CBCL or YSR.* |
| *Geibel et al. (2016) | Adolescents (aged 15-18 years) from Addis Ababa<br>*N* = 134 | YSR (2001) | Amharic | • Translation<br>• Back-translation<br>• Reviewed by expert panel<br>• Pilot test | Evaluated the psychometric properties of the YSR as a mental health screening tool for young Ethiopians vulnerable to HIV. | <u>IC (Cronbach's $\alpha$), TRR, criterion-related validity (ROC)</u><br>*See Table 5.* |
| *<sup>b</sup>Hall et al. (2014) | Somali children (mean age 11.02 years, *SD* = 2.90) living in refugee camps in Ethiopia<br>*N* = 147 | CBCL/6-18 and YSR (2001) | Somali | • Qualitative study to elicit local terms describing mental health problems<br>• Translation<br>• Back-translation<br>• Sex-related items were removed based on feedback from the community | Established psychometric properties of instruments measuring mental health problems in Somali children and adolescents living in refugee camps. | <u>IC (Cronbach's $\alpha$), combined TRR and IRR (Pearson's *r*), criterion-related validity (ROC)</u><br>*See Tables 4 and 5.* |
| <sup>#</sup>Isaksson et al. (2017) | Children (mean age = 10.3 years, *SD* = 0.43) of women living in the Butajira Rural Health Program (BRHP) area<br>*N* = 358 | CBCL/6-18 (2001) | Amharic | • Obtained an already-translated version<br>• The authors made slight modifications to the translation to make it more suited for a rural community. | Investigated a possible relationship between maternal perinatal stressors and child emotional and behavioural functioning. | <u>IC (Cronbach's $\alpha$)</u><br>Total Problems [$\alpha$ = 0.93] |
| *<sup>a</sup>Ivanova, Dobrean, et al. (2007) | Children/adolescents (aged 11-18 years) from a regional school sample (Mulatu, 1997)<br>*N* = 677 | CBCL/4-18 (1991) | Amharic | • Translation<br>• Back-translation<br>• Pilot test (*N* = 20)<br>Extracted from Mulatu (1997) | Tested the eight-syndrome structure of the CBCL across 30 societies. | <u>CFA</u><br>*See Table 4.* |
| *<sup>a</sup>Ivanova, Achenbach, et al. (2007) | Children/adolescents (aged 11-18 years) from a regional school sample (Mulatu, 1997)<br>*N* = 674 | YSR (1991) | Amharic | • Translation<br>• Back-translation<br>• Pilot test (*N* = 20)<br>Extracted from Mulatu (1997) | Tested the eight-syndrome structure of the YSR in 23 countries. | <u>CFA</u><br>*See Table 5.* |
| <sup>b</sup>Murray et al. (2018) | Children (mean age = 11.21 years, *SD* = 3.17), living in one of three refugee camps in the Somali region of Ethiopia<br>*N* = 37 | CBCL/6-18 and YSR (2001) | Somali | See Hall *et al.* (2014) | Evaluated a common elements treatment approach developed for mental illness/distress among children in Somali refugee camps. | <u>IC (Cronbach's $\alpha$)</u><br>*CBCL*<br>Internalising Problems [$\alpha$ = 0.95]<br>Externalising Problems [$\alpha$ = 0.89]<br>*YSR*<br>Internalising Problems [$\alpha$ = 0.88]<br>Externalising Problems [$\alpha$ = 0.93] |
| Uganda (*n* = 6) |  |  |  |  |  |  |
| Amone-P'Olak et al. (2007) | Adolescents (aged 14-18 years) who were formerly abducted in Northern Uganda<br>*N* = 294 | YSR (1991) | NR | NR | Investigated the prevalence of war experiences and the use of cognitive emotion regulation in response to these experiences. | <u>IC (Cronbach's $\alpha$)</u><br>Internalising Problems [$\alpha$ = 0.61]<br>Externalising Problems [$\alpha$ = 0.61] |
| *Bangirana et al. (2009) | Child survivors of cerebral malaria (aged 7-16 years)<br>*N* = 64 | CBCL/6-18 (2001) | Luganda | • Translation<br>• Back-translation | Investigated the psychometric properties of the CBCL in the context of a controlled trial aimed at improving cognitive functioning following cerebral malaria. | <u>IC (Cronbach's $\alpha$), test-retest reliability (Pearson's *r*)</u><br>*See Table 4.* |
| Eggum et al. (2011) | Children (aged 9-20 years) residing near Tororo, Uganda<br>*N* = 57 | YSR (1991) | English | • Reviewed by expert panel for cultural appropriateness<br>• Item "set fires" was removed | Determined prevalence of negative life effects and examined associations between negative life events and adjustment in Ugandan youth. | <u>IC (Cronbach's $\alpha$)</u><br>Internalising Problems (20 items) [$\alpha$ = 0.75]<br>Externalising Problems (27 items) [$\alpha$ = 0.83]<br>*Some items were not administered because they were not culturally appropriate.* |
| #Familiar et al. (2015) | HIV-infected children (aged 5-12 years, *n* = 144) and children with a history of severe malaria (aged 5-12 years, *n* = 106) | CBCL/4-18 (1991) | Luganda | • Used a previously translated version from Bangirana *et al.* (2009) | Assessed structural overlap between the CBCL and the Behaviour Rating Inventory of Executive Function (BRIEF) in a sample of Ugandan children. | <u>IC (Cronbach's $\alpha$)</u><br>*Total Problems*<br>Malaria group [$\alpha$ = 0.93]<br>HIV-infected group [$\alpha$ = 0.90] |
| Klasen et al. (2010) | Former child soldiers (aged 11-17 years)<br>*N* = 330 | YSR (2001) | Luo | • Translation<br>• Back-translation<br>• Reviewed by expert panel | Examined the effects of war and domestic violence on the mental health of child soldiers. | <u>IC (Cronbach's $\alpha$)</u><br>Alpha coefficients ranged were between 0.70 and 0.95, except for Withdrawn/Depressed syndrome scale [$\alpha$ = 0.60] and Attention Problems syndrome scale [$\alpha$ = 0.64]. |
| Ruiseñor-Escudero et al. (2015) | HIV-infected children (aged 5-12 years) living in Kayunga<br>*N* = 144 | CBCL/4-18 (1991) | Luganda | • Translation<br>• Back-translation | Investigated the association between immunological parameters and behavioural problems among children living with HIV. | <u>IC (Cronbach's $\alpha$)</u><br>Total Problems [$\alpha$ = 0.76] |
| Rwanda (*n* = 2) |  |  |  |  |  |  |
| Ng et al. (2015) | Children (aged 10-17 years) living with HIV, children with a parent living with HIV, and children unaffected by HIV, living in Kayonza or Kirehe Districts<br>*N* = 683 | YSR (1991) | Kinyarwanda | • Translation | Investigated suicidal ideation and behaviours in children living with HIV compared to children with a parent living with HIV, and children unaffected by HIV. | <u>IC (Cronbach's $\alpha$)</u><br>Internalising Problems (16 items) [$\alpha$ = 0.89] |
| Nsabimana et al. (2019) | Institutionalised and non-institutionalised children (aged 9-16 years) from rural and urban areas in Rwanda<br>*N* = 178 | CBCL/6-18 (2001) | Kinyarwanda | • Translation<br>• Back-translation<br>• Reviewed by expert panel | Explored the effects of institutionalization on externalising and internalising problems in children/adolescents. | <u>IC (Cronbach's $\alpha$)</u><br>Internalising Problems [$\alpha$ = 0.84]<br>Externalising Problems [$\alpha$ = 0.87] |
| Zambia (*n* = 1) |  |  |  |  |  |  |
| *Murray et al. (2020) | Orphaned or otherwise vulnerable adolescents (aged 13-17 years) living in Lusaka<br>*N* = 210 | YSR (2001) | English and Nyanja | • Translation<br>• Back-translation<br>• Pilot test | Evaluated the psychometric properties of various measures for assessing psychological well-being. | <u>CFA, IC (Cronbach's $\alpha$), TRR ($\rho$), criterion validity (ROC), hypothesis testing for construct validity (*W*)</u><br>*See Table 5.* |
| Central Africa ($n = 1$) | | | | | | |
| Cameroon ($n = 1$) | | | | | | |
| Wadji et al. (2020) | Children (aged 2-18 years) exposed to intimate partner violence<br>$N = 74$ | CBCL/4-18 (1991) | French and English | Used an already-translated French version | Investigated the effects of maternal childhood abuse on child psychopathology. | <u>IC (Cronbach's $\alpha$)</u><br>Internalising Problems [$\alpha = 0.84$]<br>Externalising Problems [$\alpha = 0.89$] |
| Immigrant samples ($n = 3$) | | | | | | |
| <sup>a</sup>Anakwenze & Rasmussen (2021);<br><sup>a</sup>Rasmussen et al. (2018) | Children (aged 5-12 years) with an immigrant parent from West Africa (Guinea, Sierra Leone, Senegal) who had been living in the USA for at least three years<br>$N = 91$ (2021)<br>$N = 93$ (2018) | CBCL/4-18 (1991) | Fulani and English | Translation<br>Met with interviewers during data collection to address issues of translation and wording | (2021) Investigated the impact of parental trauma, parenting difficulty, and family separation on child behaviour.<br>(2018) Investigated associations between migration and immigrant parents' perceptions of their children's safety. | <u>IC (Cronbach's $\alpha$)</u><br>Externalising Problems (32 items) [$\alpha = 0.90$] |
| <sup>#</sup>Osman et al. (2017) | Children (aged 11-16 years) with Somali-born parents, living in Sweden<br>$N = 109$ | CBCL/6-18 (2001) | Somali | Obtained permission to translate the tool from tool developer<br>Translation<br>Back-translation<br>Pilot test | Evaluated a parenting support intervention for Somali-born parents and determined its effectiveness in reducing child behavioural problems. | <u>IC (Cronbach's $\alpha$)</u><br>Cronbach's alpha coefficients for all syndrome scales were $> 0.70$.<br>*Sex-related items (59, 60, 73, 96, and 110) were excluded from the questionnaire.* |

*Note.* ASEBA = Achenbach System of Empirically Based Assessment, CBCL = Child Behaviour Checklist, YSR = Youth Self-Report, CFA = confirmatory factor analysis, IC = internal consistency, TRR = test-retest reliability, IRR = inter-rater reliability, ROC = receive operating characteristic, NR = not reported. Names of language and ethnic groups are reported verbatim from the articles. The same language is sometimes referred to by slightly different names (e.g., Xhosa = isiXhosa). The syndrome scale name 'Delinquent Behaviour/Delinquency' was updated to 'Rule-Breaking Behaviour' in the 2000/2001 versions. For journal articles corresponding to the same umbrella study: If the sample sizes and the psychometric statistics were markedly different, these articles were listed separately in different rows. If these differences were negligible, these articles are listed in a single row.
Articles marked with an asterisk (\*) are psychometric studies (i.e., the primary focus of the study was the psychometric properties of the ASEBA forms).
<sup>#</sup>Corresponding author provided clarification and/or additional information upon request.
<sup>a</sup>Multi-nation studies.
<sup>b</sup>Sample comprises refugees from other sub-Saharan African countries.

#### ASEBA Versions

Studies were published between the years 2003 and 2021. Most studies used the most recent versions of the ASEBA forms published in 2000 and 2001 (*n* = 27, 47%) or older versions published in the 1990s (*n* = 29, 50%). One study used the 2001 CBCL/6-18 and the 1991 YSR. Three studies (5%) used versions published in the 1980s. The parent report CBCL forms were used more frequently (*n* = 38, 66%) than the YSR (*n* = 28, 48%). Eight studies (14%) administered both the CBCL and the YSR. No studies used the TRF.

#### Sample Characteristics

The majority of samples comprised school-aged children and adolescents, with only four studies (7%) including pre-school aged children. Participants across all regions were typically from poorly resourced communities or populations with specific vulnerabilities (e.g., refugees, orphans, survivors of trauma or violence, living with an HIV-infected parent, etc). Although most samples were drawn from communities or schools (i.e., non-clinical populations), six samples (10%) comprised children with one or more illnesses, the most common being HIV and cerebral malaria.

Sample sizes ranged from 17 (Gershoff et al., 2010) to 3516 (Meinck et al., 2019). The average sample size across all 58 studies was 493.16 (*SD =* 642.77) and the median sample size was 281 (*IQR* = 105.25 – 600).

#### Use of the ASEBA Forms

The CBCL and YSR were primarily used as an outcome measure of child behavioural and emotional problems. Two studies, one each from South Africa and Kenya, used the CBCL and YSR respectively to estimate the prevalence of child behavioural and emotional problems (Cortina et al., 2013; Magai et al., 2018). Twenty-nine studies (50%) used one or both of the broad band scales, ‘Internalising Problems’ and ‘Externalising Problems’, and/or all problem items as a single scale (‘Total Problems’, *n* = 16, 28%). Twenty-four studies (41%) used one or more of the syndrome scales individually (i.e., not as part of a broadband scale). Only four studies (7%) used one or more of the DSM-oriented scales.

Of the 58 studies, only nine (16%) were “psychometric” studies (i.e., where a primary focus of the study was the psychometric properties of the CBCL or YSR). Interestingly, although half of the studies were from South Africa, all psychometric studies came from East African countries, namely Ethiopia (*n =* 4), Kenya (*n* = 2), Uganda (*n* = 2), and Zambia (*n* = 1).

#### Languages of Administration, Translations, and Adaptations

Forty-nine studies (84%) used at least one translated version of the tool, while five studies (9%) administered the forms exclusively in English. Information regarding the language(s) of administration was not available for two studies. In South Africa, 10 out of 29 studies (34%) administered the tool in more than one language. Studies from other regions of SSA typically administered the ASEBA forms in one of the local languages (e.g., Swahili or Luo in Kenya). Two Kenya-based studies obtained official Swahili translations from ASEBA (Magai et al., 2018, 2021). At least one study from Uganda used the Luganda translation of the CBCL prepared by Bangirana and colleagues (2009). The study from Cameroon used an existing French translation of the CBCL/4-18 from another study (Wadji et al., 2020). One study from Ethiopia also used an existing Amharic translation of the CBCL/6-18 but slightly modified some translated items to improve their comprehensibility for a rural setting (Isaksson et al., 2017). Seven studies, one from Sweden (with Somali immigrants), two from Kenya, and four from the same umbrella study in South Africa, obtained a translation license from ASEBA.

The level of detail reported about the translation and adaptation processes varied considerably. Some studies included statements such as “all research materials were translated and back-translated”. Others reported the translation process in great detail, including the number of people involved at each stage of the process, as well as each person’s qualifications and areas of expertise. Published guidelines on how to approach translation and adaptation of tools vary somewhat but tend to have overlapping features (Sousa & Rojjanasrirat, 2011). According to COSMIN translation guidelines, a ‘very good’ translation process requires (i) at least two independent forward translators with a mother-tongue in the target language, one with expertise in the construct being measured, the other naïve on the construct being measured, (ii) at least two independent back-translators, naïve on the construct being measured, with a mother tongue in the source (original) language, (iii) a clear description of how discrepancies will be resolved, (iv) a review committee (excluding the translators, preferably including the tool developer), (v) a pilot study (e.g., cognitive interview) inspecting the content validity of the translated version with a sample representative of the target population, and (vi) a written feedback report on the translation process (Mokkink et al., 2019). In light of the inconsistent reporting of the translation processes in the included studies, we could not evaluate the translations using these guidelines.

All but three studies that conducted their own translations and adaptations (*n* = 44) reported using forward and back-translation methods. Thirteen studies reported using an expert panel – typically including cultural advisors, community representatives, local healthcare workers or mental health experts, and psychometricians – to evaluate the tool instructions, response format, and items for conceptual equivalence and cultural appropriateness. Three studies also conducted interviews and focus groups with members of the target population to rate the clarity of the instructions, response format, and individual items. Fourteen studies piloted the translated versions in samples ranging in size from 20 to 200 individuals. Most of these studies did not report detailed findings of the pilot testing or focus groups. However, a few studies removed some items based on community feedback. A study from Uganda did not administer “culturally inappropriate” items on the YSR, including “I set fires” (Eggum et al., 2011). Another two South African studies from the same umbrella study removed the item “sets fires” from the CBCL (LeCroix et al., 2020; Palin et al., 2009). Interestingly, one study each from Kenya and Ethiopia also removed this item post-hoc from the YSR and CBCL respectively, as it did not perform well in confirmatory factor analyses (Ivanova, Dobrean, et al., 2007; Magai et al., 2018). A study from Ethiopia removed items related to suicide and another two studies with Somali participants removed sex-related items (Hall et al., 2014; L.K. Murray et al., 2018; Osman et al., 2017).

### Psychometric Properties

Fifty-six out of 58 studies (97%) reported internal consistency for one or more subscale using coefficient alpha (also known as Cronbach’s alpha). There was substantial variation in the alpha coefficients reported for the same subscale. For example, alpha for the CBCL Internalising Problems scale across 16 studies ranged from 0.66 to 0.95 and for the same subscale on the YSR across nine studies from 0.61 to 0.95. There were too few studies in each country and language category to conduct a stratified reliability generalisation meta-analysis. Hence, we were not able to calculate an ‘aggregated’ internal consistency statistic for each translated version of the subscales.

Among the South African studies (*n =* 29), all but two reported only internal consistency statistic(s) for one or more subscales. Eleven studies from South Africa administered the tool in more than one language, but only three of those studies reported separate alpha statistics for the different translated versions. Two studies conducted separate CFAs for the Internalising Problems and Externalising Problems broad band scales but did not report detailed results for these analyses. All studies from Kenya (*n* = 10) reported coefficient alpha statistics, three conducted CFAs, and one study reported test-retest reliability for the broad band scales, syndrome scales, and DSM-oriented scales. Studies from Ethiopia (*n* = 7) conducted more comprehensive psychometric analyses. Two studies (both by Ivanova et al., 2007 using data from Mulatu, 1997) conducted CFAs on the CBCL/4-18 and YSR respectively. Two studies evaluated criterion-related validity using receiver operating characteristic (ROC) curves. Finally, one investigated the test-retest reliability of the YSR, and another investigated combined test-retest and inter-rater reliability of both the CBCL/6-18 and the YSR. All studies from Uganda (*n* = 6) reported coefficient alpha and one study also reported test-retest reliability. The only study from Zambia conducted a comprehensive psychometric evaluation of the YSR, including CFA, internal consistency, criterion validity, test-retest reliability, and hypothesis testing. Studies from Tanzania (*n* = 1), Rwanda (*n* = 1), Cameroon (*n* = 1), and those from outside SSA with immigrant samples (*n* = 3) reported coefficient alpha only.

### COSMIN Evaluation of the Psychometric-Focused Studies

We thoroughly reviewed eight of the nine psychometric-focused studies from East African countries using COSMIN guidelines. We excluded one study from Kenya, as it was the only study that used the CBCL/1.5-5 (Kariuki et al., 2016). Four of the eight studies evaluated the School-Age version of the CBCL, and six evaluated the YSR (two studies administered both). We decided to limit the COSMIN evaluation to psychometric-focused studies only, as the remaining studies (*n* = 49) reported only internal consistency statistics, typically for one or two subscales.

Table 2 displays the COSMIN criteria for good measurement properties, with a visual icon allocated to each rating. We added a fourth rating, ‘mixed results’, to indicate a measurement property with different ratings for different sub-groups of participants. The table lists only the measurement properties and criteria that were relevant to the included studies. In this review, there were no published studies that specifically evaluated the content validity, cross-cultural validity, measurement error, or responsiveness of the ASEBA forms.

**Table 2.**
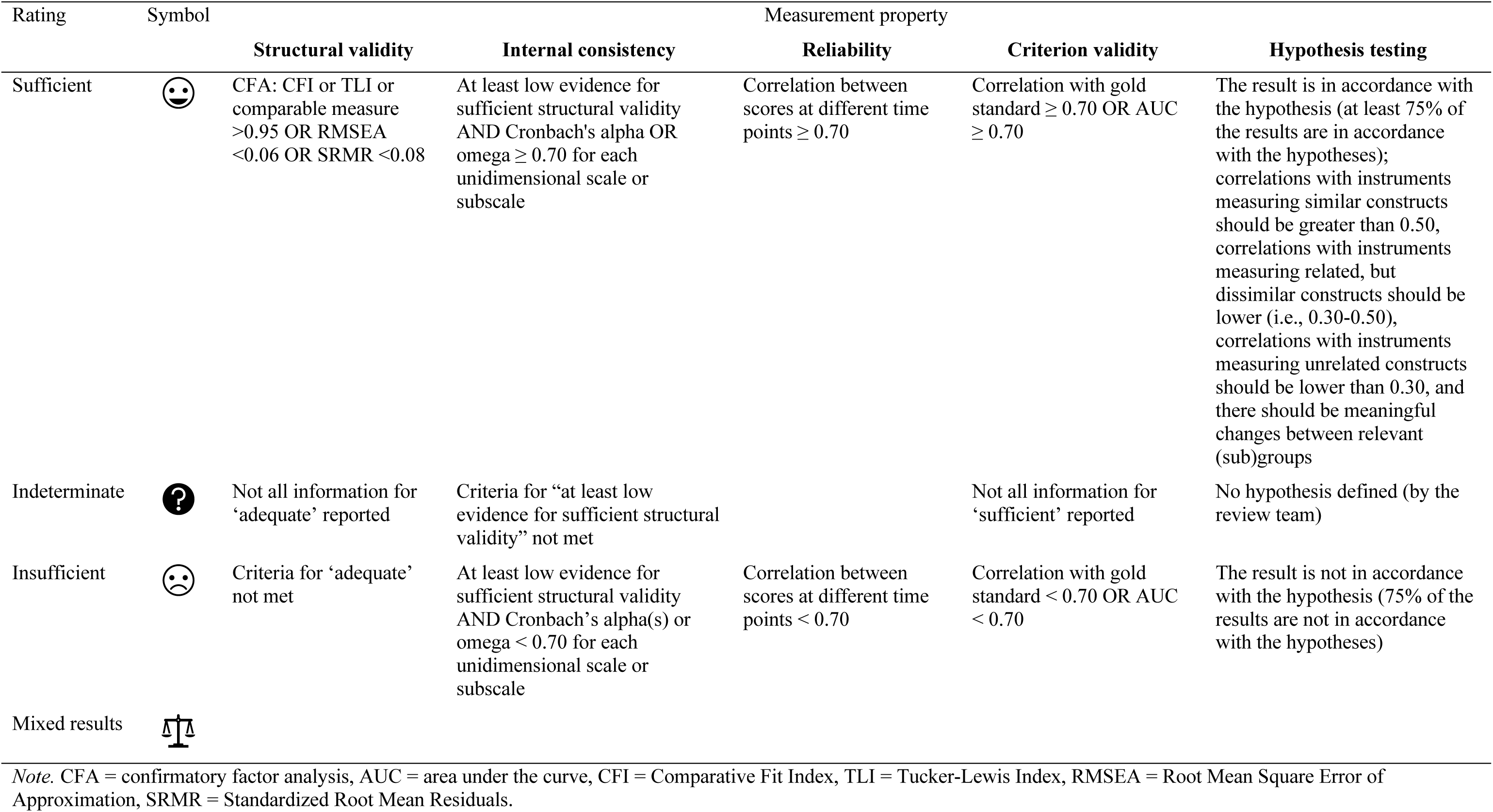
Adapted COSMIN Criteria for Adequacy of Measurement Properties

| Rating | Symbol | Measurement property |  |  |  |  |
| --- | --- | --- | --- | --- | --- | --- |
|  |  | Structural validity | Internal consistency | Reliability | Criterion validity | Hypothesis testing |
| Sufficient | | CFA: CFI or TLI or comparable measure $>0.95$ OR RMSEA $<0.06$ OR SRMR $<0.08$ | At least low evidence for sufficient structural validity AND Cronbach's alpha OR omega $\geq 0.70$ for each unidimensional scale or subscale | Correlation between scores at different time points $\geq 0.70$ | Correlation with gold standard $\geq 0.70$ OR AUC $\geq 0.70$ | The result is in accordance with the hypothesis (at least 75% of the results are in accordance with the hypotheses); correlations with instruments measuring similar constructs should be greater than 0.50, correlations with instruments measuring related, but dissimilar constructs should be lower (i.e., 0.30-0.50), correlations with instruments measuring unrelated constructs should be lower than 0.30, and there should be meaningful changes between relevant (sub)groups |
| Indeterminate |  | Not all information for 'adequate' reported | Criteria for "at least low evidence for sufficient structural validity" not met |  | Not all information for 'sufficient' reported | No hypothesis defined (by the review team) |
| Insufficient | | Criteria for 'adequate' not met | At least low evidence for sufficient structural validity AND Cronbach's alpha(s) or omega $< 0.70$ for each unidimensional scale or subscale | Correlation between scores at different time points $< 0.70$ | Correlation with gold standard $< 0.70$ OR AUC $< 0.70$ | The result is not in accordance with the hypothesis (75% of the results are not in accordance with the hypotheses) |
| Mixed results |  |  |  |  |  |  |

*Note.* CFA = confirmatory factor analysis, AUC = area under the curve, CFI = Comparative Fit Index, TLI = Tucker-Lewis Index, RMSEA = Root Mean Square Error of Approximation, SRMR = Standardized Root Mean Residuals.

We relaxed three standards set out by COSMIN. First, the guidelines state that reliability for ordinal scales should be estimated using weighted Cohen’s Kappa (*k*). However, as no studies reported this statistic, we accepted Pearson’s or Spearman’s correlation coefficients as an acceptable method to estimate reliability. Second, with regards to hypothesis testing for construct validity, COSMIN recommends that reviewers formulate a set of hypotheses about the expected magnitude and direction of the correlations between measures and mean differences in scores between groups, based on theoretical understandings of the construct, prior to the review. This is intended to reduce the possibility of bias and to ensure standardisation across studies. This was not feasible for the present review as child behaviour is such a broad construct with many possible correlates. Hence, we accepted hypotheses as long as the authors provided evidence to substantiate them. Third, for the internal consistency standard, COSMIN also requires “at least low evidence for sufficient structural validity”. However, we removed this standard for individual studies as the ASEBA forms are well-established worldwide and many studies have confirmed their structural validity (Ivanova, Achenbach, et al., 2007; Ivanova, Dobrean, et al., 2007). COSMIN also recommends that reviewers determine a reasonable “gold standard” prior to assessing the methodological quality of criterion-validity studies. We decided that a clinical assessment by a qualified mental health professional (e.g., registered clinical psychologist, psychiatrist, social worker) based on a standardised and validated diagnostic tool (e.g., DSM-5), could be considered a gold standard in this context. Table 3 displays the COSMIN criteria for evaluating the methodological quality of studies reporting psychometric properties. We assigned a colour to each rating for ease of reading. Table 4 and Table 5 present the combined results of the COSMIN analysis.

**Table 3.** COSMIN Risk of Bias Checklist for Evaluating Methodological Standards of Measurement Properties

| Rating | Measurement property |  |  |  |  |
| --- | --- | --- | --- | --- | --- |
|  | Structural validity | Internal consistency | Reliability | Criterion validity<sup>b</sup> | Hypothesis testing |
| Very good | CFA performed; *N* is 7 times the number of items and $\geq 100$; no other important methodological flaws (e.g., inappropriate estimation method) | Internal consistency statistic calculated for each unidimensional scale or subscale; Cronbach's alpha or omega calculated | Evidence provided that participants were stable in the interim period on the construct being measured; time interval appropriate<sup>a</sup>; test conditions were similar (evidence provided); Pearson/Spearman correlation coefficient calculated; no other important methodological flaws | AUC calculated OR sensitivity and specificity calculated; no other important methodological flaws | Constructs measured by comparator instrument is clear; sufficient measurement properties of comparator instrument in a population similar to study population OR adequate description of the important characteristics of the subgroups; statistical method applied was appropriate |
| Adequate | EFA performed; *N* is at least 5 times the number of items and $\geq 100$ OR at least 6 times number of items but $< 100$ | | Assumable that patients were stable; assumable that test conditions were similar | | Sufficient measurement properties of comparator instrument but unsure if these apply to study population OR adequate description of most of the important characteristics of the subgroups; assumable that statistical method was appropriate |
| Doubtful | *N* is 5 times the number of items, but $< 100$; Other minor methodological flaws | Unclear whether scale or subscale is unidimensional; only item-total correlations calculated | Unclear if patients were stable; doubtful whether time interval was appropriate or time interval was not stated; unclear if test conditions were similar; other minor methodological flaws | Other minor methodological flaws | Some information on measurement properties of the comparator instrument in any study population OR poor/no description of the important characteristics of the subgroups; statistical method applied NOT optimal |
| Inadequate | No EFA or CFA performed; *N* is $< 5$ times the number of items; Other important methodological flaws | Internal consistency statistic NOT calculated for each unidimensional scale or subscale; no Cronbach's alpha / omega and no item-total correlations calculated | Patients were NOT stable; test conditions were NOT similar; other important methodological flaws | AUC NOT calculated OR sensitivity and specificity NOT calculated; other important methodological flaws | Constructs measured by comparator instrument unclear; no information on the measurement properties of the comparator instrument(s) OR evidence for insufficient measurement properties of the comparator instrument; statistical method applied NOT appropriate |

*Note.* CFA = confirmatory factor analysis, EFA = exploratory factor analysis, AUC = area under the curve.
<sup>a</sup>COSMIN recommends an interval period of two weeks.
<sup>b</sup>We decided that a clinical assessment by a qualified mental health professional (e.g., registered clinical psychologist, psychiatrist, social worker) based on a standardised and validated diagnostic tool (e.g., DSM-5), could be considered a gold standard in this context.

**Table 4.** Measurement Properties of the CBCL/6-18 and Risk of Bias Analysis from Four Psychometric Studies Based on COSMIN Guidelines

| Measurement Property | N | Country | Broad band scales |  |  | Syndrome Scales |  |  |  |  |  |  |  | DSM-Oriented Scales |  |  |  |  |  |
| --- | --- | --- | --- | --- | --- | --- | --- | --- | --- | --- | --- | --- | --- | --- | --- | --- | --- | --- | --- |
|  |  |  | TOP | IP | EP | AB | AD | AP | RB | SP | SC | TP | WD | AFP | ANP | SOP | ADP | ODP | COP |
| Structural validity |  |  |  |  |  |  |  |  |  |  |  |  |  |  |  |  |  |  |  |
| Ivanova, Dobrean, et al. (2007) | 677 | Ethiopia |  |  |  | 😊 | 😊 | 😊 | 😊 | 😊 | 😊 | 😊 | 😊 |  |  |  |  |  |  |
| Magai et al. (2018) | 1022 | Kenya |  |  |  | 😊 | 😊 | 😊 | 😊 | 😊 | 😊 | 😊 | 😊 |  |  |  |  |  |  |
| Internal consistency |  |  |  |  |  |  |  |  |  |  |  |  |  |  |  |  |  |  |  |
| Bangirana et al. (2009) | 64 | Uganda | 😊 | 😞 | 😊 | 😊 | 😞 | 😞 | 😞 | 😞 | 😞 | 😞 | 😞 | 😞 | 😞 | 😞 | 😞 | 😞 | 😞 |
| Hall et al. (2014) | 147 | Ethiopia |  | 😊 | 😊 | 😊 | 😊 | 😊 | 😊 | 😊 | 😊 | 😊 | 😞 |  |  |  |  |  |  |
| Magai et al. (2018) | 1022 | Kenya | 😊 | 😊 | 😊 | 😊 | ⚖️ | 😊 | 😞 | 😞 | 😊 | ⚖️ | 😞 |  |  |  |  |  |  |
| Reliability |  |  |  |  |  |  |  |  |  |  |  |  |  |  |  |  |  |  |  |
| Bangirana et al. (2009) | 22 | Uganda | 😞 | 😞 | 😊 | 😊 | 😞 | 😞 | 😞 | 😊 | 😊 | 😞 | 😊 | 😞 | 😞 | 😞 | 😞 | 😊 | 😞 |
| Hall et al. (2014) | 17 | Ethiopia |  | 😞 | 😞 | 😞 | 😞 | 😞 | 😞 | 😞 | 😞 | 😊 | 😞 |  |  |  |  |  |  |
| Criterion validity |  |  |  |  |  |  |  |  |  |  |  |  |  |  |  |  |  |  |  |
| Hall et al. (2014) | 79 | Ethiopia |  | 😊 |  |  |  |  |  |  |  |  |  |  |  |  |  |  |  |
| Hypothesis testing |  |  |  |  |  |  |  |  |  |  |  |  |  |  |  |  |  |  |  |
| Hall et al. (2014) | 174 | Ethiopia |  | 😊 |  | 😞 | 😊 | ? | 😞 | ? | 😊 | ? | 😊 |  |  |  |  |  |  |

*Note.* CBCL/6-18 = Child Behaviour Checklist for Ages 6-18. TOP = Total Problems, IP = Internalising Problems, EP = Externalising Problems, AB = Aggressive Behaviour, AD = Anxious/Depressed, AP = Attention Problems, RB = Rule-breaking Behaviour, SP = Social Problems, SC = Somatic Complaints, TP = Thought Problems, WD = Withdrawn/Depressed, AFP = Affective Problems, ANP = Anxiety Problems, SOP = Somatic Problems, ADP = Attention-Deficit/Hyperactive Problems, ODP = Oppositional Defiant Problems, COP = Conduct Problems. For more information about the Risk of Bias ratings for individual studies, please contact the corresponding author.

**Table 5.**
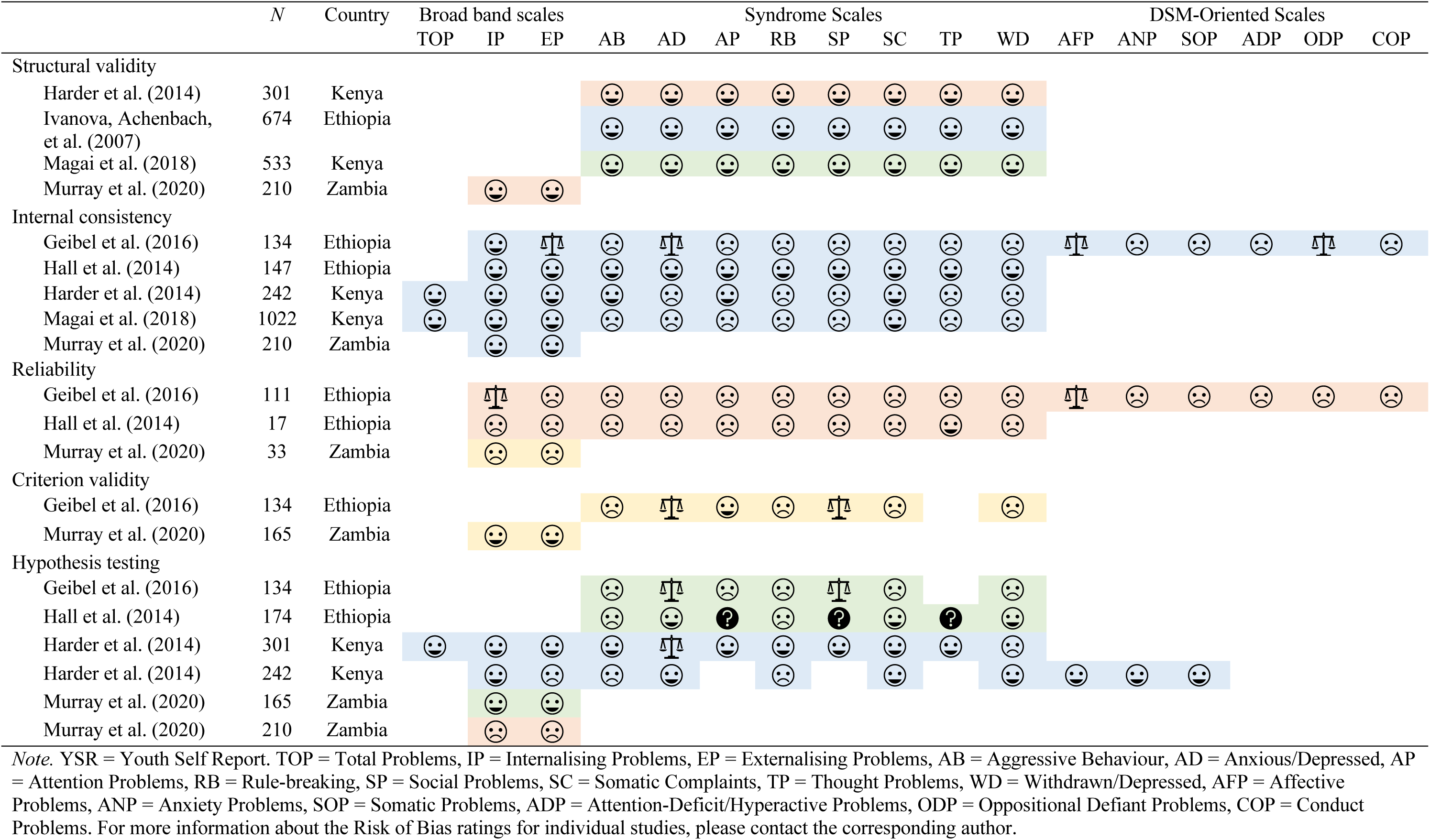
Measurement Properties of the YSR and Risk of Bias Analysis from Six Psychometric Studies Based on COSMIN Guidelines

All studies measuring structural validity conducted a CFA using tetrachoric or polychoric correlation matrices with a robust weighted least squares estimator, which is recommended for ordinal data (Li, 2016). In terms of structural validity, two ‘very good’ quality studies, one each from Kenya and Ethiopia, supported the factorial structure of the CBCL syndrome scales using CFAs. The YSR syndrome scales also performed well in CFAs across all four studies, although the methodological qualities of three studies were somewhat compromised by the smaller sample sizes. These findings suggest that the latent constructs measured by the ASEBA syndrome scales are being adequately explained by the specific behavioural problems (i.e., items) in these populations (De Kock et al., 2013).

Internal consistency was the most commonly reported measurement property, with three and five studies reporting coefficient alpha for CBCL and YSR subscales respectively. The COSMIN methodological standards for internal consistency are minimal, hence the overall quality of the methods used for this measurement property were ‘very good’ overall. Alpha coefficients for the broadband scales were generally higher than those for the syndrome scales and DSM-oriented scales. This was not surprising as the value of alpha is influenced by the length of the tool (Cortina, 1993). For the CBCL, the Aggressive Behaviour, Attention Problems, and Somatic Complaints syndrome scales performed the best in terms of internal consistency, meeting the ‘sufficient’ criteria in at least two out of the three studies. The Withdrawn/Depressed syndrome scale, however, did not meet the necessary criteria in any of the three studies. One study in this category had a relatively smaller sample size (*n* = 64; Bangirana et al., 2009) than the other two studies measuring internal consistency of the CBCL. In this smaller study, all but one of the syndrome and DSM-oriented scales did not meet the ‘sufficient’ criteria. For the YSR, Somatic Complaints was the only syndrome scale to meet the criteria in at least three of the four studies. Only two studies (one each for the CBCL and YSR) estimated internal consistencies for the DSM-oriented subscales, and these results were consistently insufficient.

The methodological quality of the reliability analyses for both the CBCL and the YSR were either ‘doubtful’ or ‘inadequate’. Reasons for the poorer quality of studies were too long or too short time intervals between administrations (e.g., 9 weeks, 5-7 days), participants undergoing an intervention in between administrations, and other methodological flaws, including not specifying how a subset of the sample was selected for re-administration, and a lack of evidence that participants were stable on the construct to be measured in between administrations. In terms of the measurement property itself, the correlation coefficients were consistently insufficient across forms and subscales. Overall, current evidence for test-retest reliability of the ASEBA forms is inadequate in these SSAn populations.

In terms of criterion validity, Geibel and colleagues’ (2016) study was the only one to use a psychiatric assessment as a gold standard for the ROC analysis. However, these assessments were not based on a standardized clinical diagnostic tool. Two studies developed their own criteria to identify cases (Hall et al., 2014; S.M. Murray et al., 2020). Murray and colleagues (2020) created a four-item screening questionnaire, administered to the child and their caregiver, asking whether the child had significant psychosocial problems (“yes” or “no”). In Hall and colleagues’ (2014) study, refugee camp social workers identified cases using a list of common internalising and externalising symptoms. The social workers’ assessments were then corroborated with caregivers’ responses to a short screening questionnaire. Based on the few studies included in this analysis, there is very limited evidence to substantiate the criterion validity of the ASEBA forms in SSA.

Four studies conducted some form of hypothesis testing to examine the construct validity of the ASEBA subscales. All four studies evaluated known-groups validity based on clinical characteristics of the sample (i.e., ‘case’ vs ‘non-case’) with mixed results, and one study from Kenya examined sex differences in levels of internalising and externalising behaviours respectively (Harder et al., 2014). The study from Zambia estimated convergent and divergent validity of the Internalising Problems and Externalising Problems subscales on the YSR (S.M. Murray et al., 2020). Only two out of the five comparator measures used (measuring post-traumatic stress and well-being, respectively) had adequate psychometric properties in a similar population. For both the Internalising Problems and Externalising Problems subscales, less than 75% of the results were in accordance with the hypotheses.

## Discussion

We identified 145 studies that used the ASEBA forms to measure child behaviour problems in SSAn samples. This suggests that the ASEBA forms are used frequently, at least for research purposes, in SSAn contexts. However, less than half of the studies reported any measurement properties of the ASEBA forms. Of the studies that did report measurement properties, most reported only coefficient alpha as a measure of internal consistency for the subscales used. The widespread use of the ASEBA forms in SSA without evaluation of measurement properties warrants consideration. A tool’s measurement properties are inextricably tied to the context in which it is administered. Without sufficient evidence to support the validity of the information derived from the tools used, the dependability of results remains questionable. The tendency of applied researchers to conduct and report limited psychometric evaluations only (i.e., coefficient alpha), without any further investigation or interpretation, remains a challenge to research in this field. Comprehensive psychometric analyses are necessary to arrive at meaningful and accurate conclusions about a tool’s measurement properties (Dima, 2018). In addition, psychometric analyses should be reported clearly and comprehensively, and this information should be easily accessible to readers. COSMIN is in the process of developing a checklist for standards on reporting measurement properties (see https://www.cosmin.nl/tools/checklists-assessing-methodological-study-qualities/). This will hopefully aid in developing a standardised and transparent approach to reporting measurement properties in research studies.

Most of the studies included in the final analysis administered at least one translated version of the CBCL or YSR, and almost all translations were created specifically for use in those studies. There were inconsistencies with regards to the reporting of the translation procedures. Descriptions of the translation procedures provided little detail, raising doubts about the integrity of the translations, as judged by COSMIN standards. Although it is possible that rigorous methodological guidelines were adhered to, this information was not readily available in most cases. Consequently, we were not able to evaluate the translation procedures across studies. The quality of a translation may significantly impact the validity of a tool (De Kock et al., 2013; Van Widenfelt et al., 2005). In a sense, a translated version of a tool becomes its own outcome measure that should be evaluated for content validity (Terwee, Prinsen, Chiarotto, de Vet, et al., 2018). Although a few studies reported the use of focus groups and pilot testing to assess relevance and comprehensibility, the results of these investigations (e.g., any re-wordings or modifications made to the original draft) were not always reported. Transparent reporting of translations and adaptations serves two important purposes. First, it grants readers the opportunity to evaluate the validity of the translated versions. Second, it serves as a useful record for researchers or clinicians who may be interested in administering the translated tool in future studies or in clinical settings. In this review, we found that only three out of the ten studies that administered the ASEBA forms in more than one language conducted separate psychometric analyses for each version. This would be considered an important step to rule out potential measurement bias.

In this review, we also evaluated eight of the nine psychometric-focused studies using COSMIN standards and criteria for good measurement properties. To our knowledge, these nine studies are the only published journal articles addressing the validity of the ASEBA forms in SSAn contexts. Overall, evidence to support the validity and reliability of the CBCL and YSR in SSAn countries in the existing literature is limited. Furthermore, the variable quality of the methods used across different studies to assess the measurement properties of the CBCL and YSR preclude us from making confident recommendations regarding its use in these regions.

Having said this, the statistical methods used, as assessed by the COSMIN Risk of Bias Checklist, were generally adequate. The main exceptions to this were the reliability and criterion validity analyses. More studies with different designs and larger samples are needed to learn about the criterion validity, test-retest reliability, and inter-rater reliability of the ASEBA forms in SSA. Criterion validity is a very important measurement property if the ASEBA forms are to be used as screening tools in community settings. Although no single “gold-standard” instrument currently exists for child behavioural and emotional problems, judging ASEBA scores against clinical assessments based on standardized diagnostic tools may be a strong starting point.

Coefficient alpha was the most frequently reported statistic across all studies. However, there are limitations of coefficient alpha as a measure of internal consistency (Dunn et al., 2014; Sijtsma, 2009). Ordinal coefficient alpha may generate a more reliable estimate of internal consistency for Likert scales, such as the ASEBA forms (Zumbo et al., 2007). In terms of structural validity, the majority of studies were of a very good standard, barring a few studies with sample sizes smaller than required for a tool measuring multiple constructs with many items. Although the broadband scales (Internalising Problems and Externalising Problems) were frequently administered in SSA, no studies conducted higher order or bifactor CFAs that would have investigated the unidimensionality of these broad band scales.

Most of the studies included in the final analysis came from South Africa, although none of these studies were specifically focused on the measurement properties of the ASEBA forms. Hence, there remains limited evidence to support the validity of the ASEBA forms in a South African context. A smaller but significant proportion of the included studies came from East African countries (notably Kenya, Ethiopia, and Uganda). All nine psychometric-focused studies came from East African countries. Compared to Southern and East Africa, there were very few studies from West and Central Africa. It is possible that other measures of child and behavioural problems are more popular in these regions. Although there was only one study that came from a West African country (i.e., Ghana), individuals of West African origin living outside of SSA were also represented in the included studies.

## Limitations

The two reviewers made every effort to ensure that all papers were thoroughly screened and reviewed. However, it is possible that a few articles were either not included in the search results, accidentally removed from the reference library, or incorrectly screened. A limitation of our study was the exclusion of unpublished “grey” literature, including theses, books, and conference presentations. Although we made the decision to exclude these records for practical reasons, grey literature would have likely enriched our analysis and reduced the risk of publication bias. Another important limitation of our study was that we could not use the COSMIN GRADE approach to quantitatively pool the results from individual studies and grade the overall quality of evidence for each measurement property (Mokkink, Prinsen, et al., 2018). Results from individual studies were too inconsistent to pool quantitatively. Moreover, there were too few studies in each “sub-group” (e.g., country, language of administration, sample characteristics) to arrive at reliable conclusions for each possible combination of subscale and sub-group.

## Gaps Identified and Recommendations

One important gap in the current literature is the dearth of studies evaluating the content validity of the ASEBA forms in SSA. Content validity, the extent to which the content of a tool adequately represents the construct it measures, is arguably the most important of all measurement properties (Terwee, Prinsen, Chiarotto, Westerman, et al., 2018). If a tool does not have content validity, then all other measurement properties are irrelevant. As described earlier, there were some attempts to evaluate the relevance and comprehensibility of the ASEBA forms through pilot testing. To our knowledge, only one included study explored the comprehensiveness of the ASEBA forms. Prior to conducting their study, Hall and colleagues (2014) used qualitative methods to identify local symptoms of internalising and externalising behaviours in Somali refugees living in Ethiopia. The authors added 11 and 4 of these locally derived symptoms to the Internalising Problems and Externalising Problems subscales respectively. Although we could not include this preliminary study in the current analysis, the findings emphasise the importance of evaluating the comprehensiveness of behavioural screening tools in SSA.

## Summary and Conclusion

The primary aim of the present review was to investigate the measurement properties of the ASEBA forms in SSAn countries, where translated versions of the forms are frequently administered. At present, evidence is limited in terms of both the number and quality of available studies. East African countries have already made significant progress with regards to evaluating translated versions of the ASEBA forms in local contexts. In South Africa, however, the measurement properties of the ASEBA forms remain under-studied despite their widespread use in research. Data from other areas of SSA are largely absent. This review has demonstrated the importance of validating existing behavioural tools for culturally and linguistically diverse contexts in SSA. Comprehensive and ongoing psychometric evaluations of tools require time and resources. However, the result is that clinicians and researchers become more confident that the inferences made based on these tools are accurate and dependable.

## Supporting information

digital supplement

## Data Availability

All data produced in the present work (i.e., information extracted from published studies) are contained in the manuscript.

## Acknowledgements

We thank Mrs Mary Shelton from the University of Cape Town’s Health Sciences Library for her advice and assistance with the systematic review search strategy.

