## Supplementary material for "Psychometric Properties of the ASEBA Child Behaviour Checklist and Youth Self-Report in Sub-Saharan Africa: A Systematic Review": digital supplement

### Supplementary Materials

Table S1: *Summary of Databases, Search Terms, and Number of Studies Pulled from Each Database*
Table S2: *Excluded Studies and Reasons for Exclusion (n = 64)*
Appendix 1: *Journal articles with ASEBA Forms, a Sub-Saharan African sample, but no Psychometric Properties (n = 87)*

**Table S1**

*Summary of Databases, Search Terms, and Number of Studies Pulled from Each Database*

| Database | Search terms | Notes | Number of studies | |
|---|---|---|---|---|
| PubMed | (((“child behaviour checklist” OR “child behavior checklist” OR CBCL OR “youth self-report” OR “teacher’s report form”))<br>AND<br>(Africa OR African OR Angola OR Benin OR Botswana OR “Burkina Faso” OR Burundi OR Cameroon OR “Central African Republic” OR Chad OR Congo OR “Democratic Republic of Congo” OR “Republic of Congo” OR Djibouti OR “Equatorial Guinea” OR Eritrea OR Eswatini OR Ethiopia OR Gabon OR Gambia OR Ghana OR Guinea OR “Guinea Bissau” OR “Ivory Coast” OR “Cote d’Ivoire” OR Kenya OR Lesotho OR Liberia OR Malawi OR Mali OR Mauritania OR Mozambique OR Mocambique OR Namibia OR Niger OR Nigeria OR Rwanda OR Senegal OR “Sierra Leone” OR Somalia OR “South Africa” OR Sudan OR South Sudan OR Swaziland OR Tanzania OR Togo OR Uganda OR Zaire OR Zambia OR Zimbabwe OR “sub-Saharan Africa” OR “sub-Saharan African”))<br>NOT<br>(“African American” OR “African-American”) | “All fields” selected for all three search boxes.<br><br>No filters or restrictions were applied (e.g., date, document type, full text availability). | | 129 |
| EBSCOhost<br><br>Academic Search Premier<br>Africa-Wide Information<br>Health Source: Nursing/Academic Edition<br>CINAHL<br>ERIC<br>APA PsycInfo<br>APA PsycArticles | TX (“child behaviour checklist” OR “child behavior checklist” OR CBCL)<br>AND<br>TX (achenbach or ASEBA)<br>AND<br>TX (Africa OR Angola OR Benin OR Botswana OR “Burkina Faso” OR Burundi OR Cameroon OR “Central African Republic” OR Chad OR Congo OR “Democratic Republic of Congo” OR “Republic of Congo” OR Djibouti OR “Equatorial Guinea” OR Eritrea OR Eswatini OR Ethiopia OR Gabon OR Gambia OR Ghana OR Guinea OR “Guinea Bissau” OR “Ivory Coast” OR “Cote d’Ivoire” OR Kenya OR Lesotho OR Liberia OR Malawi OR Mali OR Mauritania OR Mozambique OR Mocambique OR Namibia OR Niger OR Nigeria OR Rwanda OR Senegal OR “Sierra Leone” OR Somalia OR “South Africa” OR Sudan OR South Sudan OR Swaziland OR Tanzania OR | “All text” fields were selected for all search boxes.<br><br>The initial search yielded too many results (1745). We narrowed the search by adding another line to the search (achenbach OR ASEBA).<br><br>No filters or restrictions were applied. | Total<br><br>Academic Search<br>Africa-Wide<br>Health Source<br>CINAHL<br>ERIC<br>APA PsycInfo<br>APA PsycArticles | 853<br><br>512<br>17<br>96<br>45<br>1<br>62<br>120 |

| | | | |
|---|---|---|---|
| | Togo OR Uganda OR Zaire OR Zambia OR Zimbabwe OR “sub-Saharan Africa” OR “sub-Saharan African”)<br>NOT<br>TX (“African American” OR “African-American”) | | |
| Scopus | ALL ("Child behaviour checklist" OR "Child behavior checklist" OR cbcl OR "youth self-report" OR "teacher's report form")<br>AND<br>ALL (achenbach OR aseba)<br>AND<br>ALL (Africa OR African OR "sub-Saharan Africa" OR "sub-Saharan AND African" OR Angola OR Benin OR Botswana OR "Burkina Faso" OR Burundi OR Cameroon OR "Central African Republic" OR chad OR Congo OR "Democratic Republic of Congo" OR "Republic of Congo" OR Djibouti OR "Equatorial Guinea" OR Eritrea OR Eswatini OR Ethiopia OR Gabon OR Gambia OR Ghana OR guinea OR "Guinea Bissau" OR "Ivory Coast" OR "Cote d'Ivoire" OR Kenya OR Lesotho OR Liberia OR Malawi OR Mali OR Mauritania OR Mozambique OR Mocambique OR Namibia OR Niger OR Nigeria OR Rwanda OR Senegal OR "Sierra Leone" OR Somalia OR "South Africa" OR Sudan OR "South Sudan" OR Swaziland OR Tanzania OR Togo OR Uganda OR Zaire OR Zambia OR Zimbabwe))<br>AND NOT<br>ALL ("African American" OR "African-American")<br>AND NOT<br>INDEX (medline) | Results from MEDLINE were filtered out to avoid duplicates of the PubMed search results. The initial search yielded too many results (1288) with the phrase “youth self-report” that were not relevant to the ASEBA forms. We therefore narrowed the search by adding (achenbach OR ASEBA).<br><br>No other filters or restrictions were applied. | 204 |
| Google Scholar | ("child behaviour checklist" OR "child behaviour checklist" OR cbcl OR “youth self-report” OR “teacher’s report form")<br>AND (Africa OR African OR "sub-Saharan Africa" OR “sub-Saharan African”) -"African American" | We included all results from the first twenty pages (20 x 10 results per page), as the results became less relevant after this point. We simplified the SSA search terms due to space limits in the search box. | 200 |
| Proquest | ("child behaviour checklist" OR "child behavior checklist" OR “youth self-report” or “teacher’s report form”)<br>AND | The initial search looked for search terms “Anywhere”, but this yielded too many results (1261). Restricting the SSA | 37 |

| | | | |
|---|---|---|---|
| | ab(Africa OR African OR Angola OR Benin OR Botswana OR "Burkina Faso" OR Burundi OR Cameroon OR "Central African Republic" OR Chad OR Congo OR "Democratic Republic of Congo" OR "Republic of Congo" OR Djibouti OR "Equatorial Guinea" OR Eritrea OR Eswatini OR Ethiopia OR Gabon OR Gambia OR Ghana OR Guinea OR "Guinea Bissau" OR "Ivory Coast" OR "Cote d'Ivoire" OR Kenya OR Lesotho OR Liberia OR Malawi OR Mali OR Mauritania OR Mozambique OR Mocambique OR Namibia OR Niger OR Nigeria OR Rwanda OR Senegal OR "Sierra Leone" OR Somalia OR "South Africa" OR Sudan OR "South Sudan" OR Swaziland OR Tanzania OR Togo OR Uganda OR Zaire OR Zambia OR Zimbabwe OR "sub-Saharan Africa" OR "sub-Saharan African")<br>NOT<br>("African American" OR "African-American") | search terms to the 'Location' search field yielded too few results (4). The final search restricted the SSA search terms to "Abstract". All search results were limited to books, book chapters, conference papers and proceedings, and thesis/dissertations. Only 1 result was a book and the rest were theses. | |
| PRIMO University of Cape Town Libraries Catalogue | ('Child behaviour checklist' OR 'child behavior checklist') AND (africa OR african OR 'sub-Saharan Africa' OR 'sub-saharan African') NOT ('african american' OR 'african-american') | Results were filtered to include books, book chapters, dissertations and conference proceedings.<br><br>The full search terms used for other databases yielded too many results (4795). We modified the SSA search terms but there were still too many results (3271). Finally, we simplified the ASEBA terms to "child behaviour checklist" OR "child behavior checklist" only. | 169 |

*Note.* CBCL = Child Behaviour Checklist, ASEBA = Achenbach System of Empirically Based Assessment, SSA = sub-Saharan Africa.

**Table S2**

*Excluded Studies and Reasons for Exclusion (n = 64)*

| Reference | Country | Reason for exclusion |
|---|---|---|
| East Africa (*n* = 20) | | |
| Aramburu Alegret, I., Pérez-Testor, C., Mercadal Rotger, J., Salamero Baró, M., Davins Pujols, M., Mirabent Junyent, V., Aznar Martínez, B., & Brodzinsky, D. (2020). Influence of communicative openness on the psychological adjustment of internationally adopted adolescents. *Journal of Research on Adolescence*, *30*, 226–237. https://doi.org/10.1111/jora.12464 | Ethiopia (Spain) | No stratified data for Ethiopian participants |
| Bjørknes, R., Manger, T., Bjørknes, R., & Manger, T. (2013). Can parent training alter parent practice and reduce conduct problems in ethnic minority children? A randomized controlled trial. *Prevention Science*, *14*(1), 52–63. https://doi.org/10.1007/s11121-012-0299-9 | Somalia (Norway) | No stratified data for Somali participants |
| Bornstein, M. H., Putnick, D. L., Lansford, J. E., Al-Hassan, S. M., Bacchini, D., Bombi, A. S., Chang, L., Deater-Deckard, K., Di Giunta, L., Dodge, K. A., Malone, P. S., Oburu, P., Pastorelli, C., Skinner, A. T., Sorbring, E., Steinberg, L., Tapanya, S., Tirado, L. M. U., Zelli, A., & Alampay, L. P. (2017). "Mixed blessings": Parental religiousness, parenting, and child adjustment in global perspective. *Journal of Child Psychology & Psychiatry*, *58*(8), 880–892. https://doi.org/10.1111/jcpp.12705 | Kenya | No stratified data for Kenyan participants |
| Cederblad, M., & Hook, B. (1999). Mental health in international adoptees as teenagers and young adults: An epidemiological study. *Journal of Child Psychology & Psychiatry & Allied Disciplines*, *40*(8), 1239. https://doi.org/10.1111/1469-7610.00540 | Ethiopia (Sweden) | No stratified data for Ethiopian participants |
| Chang, L., Lu, H. J., Lansford, J. E., Skinner, A. T., Bornstein, M. H., Steinberg, L., Dodge, K. A., Chen, B. Bin, Tian, Q., Bacchini, D., Deater-Deckard, K., Pastorelli, C., Alampay, L. P., Sorbring, E., Al-Hassan, S. M., Oburu, P., Malone, P. S., Di Giunta, L., Tirado, L. M. U., & Tapanya, S. (2019). Environmental harshness and unpredictability, life history, and social and academic behavior of adolescents in nine countries. *Developmental Psychology*, *55*(4), 890–903. https://doi.org/10.1037/dev0000655 | Kenya | No stratified data for Kenyan participants |
| Chaudhury, S., Brown, F. L., Kirk, C. M., Mukunzi, S., Nyirandagijimana, B., Mukandanga, J., Ukundineza, C., Godfrey, K., Ng, L. C., Brennan, R. T., & Betancourt, T. S. (2016). Exploring the potential of a family-based prevention intervention to reduce alcohol use and violence within HIV-affected families in Rwanda. *AIDS Care*, *28*, 118–129. https://doi.org/10.1080/09540121.2016.1176686 | Rwanda | Non-standard use |
| Deater-Deckard, K., Godwin, J., Lansford, J. E., Bacchini, D., Bombi, A. S., Bornstein, M. H., Chang, L., Di Giunta, L., Dodge, K. A., Malone, P. S., Oburu, P., Pastorelli, C., Skinner, A. T., Sorbring, E., Steinberg, L., Tapanya, S., Alampay, L. P., Uribe Tirado, L. M., Zelli, A., & Al-Hassan, S. M. (2018). Within- and between-person and group variance in behavior and beliefs in cross-cultural longitudinal data. *Journal of Adolescence*, *62*, 207–217. https://doi.org/10.1016/j.adolescence.2017.06.002 | Kenya | No stratified data for Kenyan participants |

| | | |
|---|---|---|
| Eggum-Wilkens, N. D., Zhang, L., & Farago, F. (2017). Karamojong adolescents in Tororo, Uganda: Life events, adjustment problems, and protective factors. *Journal of Loss & Trauma*, *22*(4), 283–296. https://doi.org/10.1080/15325024.2017.1284508 | Uganda | Non-standard use |
| Elovainio, M., Hakulinen, C., Pulkki-Råback, L., Raaska, H., & Lapinleimu, H. (2018). The network structure of childhood psychopathology in international adoptees. *Journal of Child & Family Studies*, *27*(7), 2161–2170. https://doi.org/10.1007/s10826-018-1046-z | Ethiopia and Mozambique (Finland) | No stratified data for Ethiopian or Mozambican participants |
| Elovainio, M., Raaska, H., Sinkkonen, J., Mäkipää, S., & Lapinleimu, H. (2015). Associations between attachment-related symptoms and later psychological problems among international adoptees: Results from the FinAdo study. *Scandinavian Journal of Psychology*, *56*(1), 53–61. https://doi.org/10.1111/sjop.12174 | Ethiopia and Mozambique (Finland) | No stratified data for Ethiopian or Mozambican participants |
| Kane, J. C., Murray, L. K., Bass, J. K., Johnson, R. M., & Bolton, P. (2016). Validation of a substance and alcohol use assessment instrument among orphans and vulnerable children in Zambia using Audio Computer Assisted Self-Interviewing (ACASI). *Drug and Alcohol Dependence*, *166*, 85–92. https://doi.org/https://doi.org/10.1016/j.drugalcdep.2016.06.026 | Zambia | Non-standard use |
| Lansford, J. E., Chang, L., Dodge, K. A., Malone, P. S., Oburu, P., Palmérus, K., Bacchini, D., Pastorelli, C., Bombi, A. S., Zelli, A., Tapanya, S., Chaudhary, N., Deater-Deckard, K., Manke, B., & Quinn, N. (2005). Physical discipline and children's adjustment: Cultural normativeness as a moderator. *Child Development*, *76*(6), 1234–1246. https://doi.org/10.1111/j.1467-8624.2005.00847.x | Kenya | No stratified ASEBA data for Kenyan participants |
| Lim, S.-L., & Ogawa, Y. (2014). "Once I had kids, now I am raising kids": Child-Parent Relationship Therapy (CPRT) with a Sudanese refugee family—A case study. *International Journal of Play Therapy*, *23*(2), 70–89. https://doi.org/10.1037/a0036362 | Sudan | Case study |
| Mulatu, M. S. (1995). Prevalence and risk factors of psychopathology in Ethiopian children. *Journal of the American Academy of Child & Adolescent Psychiatry*, *34*(1), 100–109. https://doi.org/10.1097/00004583-199501000-00020 | Ethiopia | Non-standard use |
| Nduwimana, E., Mukunzi, S., Ng, L. C., Kirk, C. M., Bizimana, J. I., & Betancourt, T. S. (2017). Mental health of children living in foster families in rural Rwanda: The role of HIV and the family environment. *AIDS and Behavior*, *21*(6), 1518–1529. https://doi.org/10.1007/s10461-016-1482-y | Rwanda | Non-standard use |
| Putnick, D. L., Bornstein, M. H., Lansford, J. E., Malone, P. S., Pastorelli, C., Skinner, A. T., Sorbring, E., Tapanya, S., Tirado, L. M. U., Zelli, A., Alampay, L. P., Al-Hassan, S. M., Bacchini, D., Bombi, A. S., Chang, L., Deater-Deckard, K., Di Giunta, L., Dodge, K. A., & Oburu, P. (2015). Perceived mother and father acceptance-rejection predict four unique aspects of child adjustment across nine countries. *Journal of Child Psychology and Psychiatry*, *56*(8), 923–932. https://doi.org/10.1111/jcpp.12366 | Kenya | No stratified data for Kenyan participants |
| Rita, N., Elovainio, M., Raaska, H., Lahti-Nuuttila, P., Matomäki, J., Sinkkonen, J., & Lapinleimu, H. (2017). Child and family-related predictors of psychological outcomes in children adopted from abroad; what is the role of caregiver time? *Scandinavian Journal of Psychology*, *58*(4), 312–317. https://doi.org/10.1111/sjop.12374 | Ethiopia and Mozambique (Finland) | No stratified data for Ethiopian or Mozambican participants |

| | | |
|---|---|---|
| Roskam, I., Stievenart, M., Tessier, R., Muntean, A., Escobar, M., Santelices, M., Juffer, F., Ijzendoorn, M., & Pierrehumbert, B. (2014). Another way of thinking about ADHD: The predictive role of early attachment deprivation in adolescents' level of symptoms. *Social Psychiatry & Psychiatric Epidemiology*, *49*(1), 133–144. https://doi.org/10.1007/s00127-013-0685-z | Ethiopia | No stratified data for Ethiopian participants |
| Schenck-Fontaine, A., Lansford, J. E., Skinner, A. T., Deater-Deckard, K., Di Giunta, L., Dodge, K. A., Oburu, P., Pastorelli, C., Sorbring, E., & Steinberg, L. (2020). Associations between perceived material deprivation, parents' discipline practices, and children's behavior problems: An international perspective. *Child Development*, *91*(1), 307–326. https://doi.org/https://doi.org/10.1111/cdev.13151 | Kenya | No stratified data for Kenyan participants |
| Vollebergh, W. A. M., Have, M., Dekovic, M., Oosterwegel, A., Pels, T., Veenstra, R., Winter, A., Ormel, H., Verhulst, F., ten Have, M., Veenstra, R., & de Winter, A. (2005). Mental health in immigrant children in the Netherlands. *Social Psychiatry & Psychiatric Epidemiology*, *40*(6), 489–496. https://doi.org/10.1007/s00127-005-0906-1 | Somalia (The Netherlands) | No stratified data for Somali participants |
| Southern Africa ($n$ = 19) | | |
| Barbarin, O. A., & Richter, L. (2001). Economic status, community danger and psychological problems among South African children. *Childhood*, *8*(1), 115–133. https://doi.org/10.1177/0907568201008001007 | South Africa | Non-standard use |
| Barone, L., Lionetti, F., & Green, J. (2017). A matter of attachment? How adoptive parents foster post-institutionalized children's social and emotional adjustment. *Attachment & Human Development*, *19*(4), 323–339. https://doi.org/10.1080/14616734.2017.1306714 | South Africa | No stratified data for South Africa participants |
| Cluver, L. D., Orkin, F. M., Campeau, L., Toska, E., Webb, D., Carlqvist, A., & Sherr, L. (2019). Improving lives by accelerating progress towards the UN Sustainable Development Goals for adolescents living with HIV: a prospective cohort study. *The Lancet Child & Adolescent Health*, *3*(4), 245–254. https://doi.org/10.1016/s2352-4642(19)30033-1 | South Africa | Non-standard use |
| Cluver, L. D., Rudgard, W. E., Toska, E., Zhou, S., Campeau, L., Shenderovich, Y., Orkin, M., Desmond, C., Butchart, A., & Taylor, H. (2020). Violence prevention accelerators for children and adolescents in South Africa: A path analysis using two pooled cohorts. *PLoS Medicine*, *17*(11), e1003383. https://doi.org/10.1371/journal.pmed.1003383 | South Africa | Non-standard use |
| Davies, L. C., & McKelvey, R. S. (1998). Emotional and behavioural problems and competencies among immigrant and non-immigrant adolescents. *Australian & New Zealand Journal of Psychiatry*, *32*(5), 658–665. https://doi.org/10.3109/00048679809113120 | South Africa | No stratified data for South Africa participants |
| de la Loge, C., Hunter, S. J., Schiemann, J., & Yang, H. (2010). Assessment of behavioral and emotional functioning using standardized instruments in children and adolescents with partial-onset seizures treated with adjunctive levetiracetam in a randomized, placebo-controlled trial. *Epilepsy & Behavior*, *18*(3), 291–298. https://doi.org/10.1016/j.yebeh.2010.04.017 | South Africa | No stratified data for South Africa participants |

| | | |
|---|---|---|
| Diliberto, R., & Kearney, C. A. (2018). Latent class symptom profiles of selective mutism: Identification and linkage to temperamental and social constructs. *Child Psychiatry and Human Development*, *49*(4), 551–562. https://doi.org/10.1007/s10578-017-0774-4 | South Africa | No stratified data for South Africa participants |
| Gardner, F., Waller, R., Maughan, B., Cluver, L., & Boyes, M. (2015). What are the risk factors for antisocial behavior among low-income youth in Cape Town? *Social Development*, *24*(4), 798–814. https://doi.org/10.1111/sode.12127 | South Africa | Non-standard use |
| Gautam, P., Lebel, C., Narr, K. L., Mattson, S. N., May, P. A., Adnams, C. M., Riley, E. P., Jones, K. L., Kan, E. C., & Sowell, E. R. (2015). Volume changes and brain-behavior relationships in white matter and subcortical gray matter in children with prenatal alcohol exposure. *Human Brain Mapping*, *36*(6), 2318–2329. https://doi.org/10.1002/hbm.22772 | South Africa | No stratified data for South Africa participants |
| Govender, K., Reardon, C., Quinlan, T., & George, G. (2014). Children's psychosocial wellbeing in the context of HIV/AIDS and poverty: a comparative investigation of orphaned and non-orphaned children living in South Africa. *BMC Public Health*, *14*(1), 998–1021. https://doi.org/10.1186/1471-2458-14-615 | South Africa | Non-standard use |
| Hoare, J., Stein, D. J., Heany, S. J., Fouche, J.-P., Phillips, N., Er, S., Myer, L., Zar, H. J., Horvath, S., & Levine, A. J. (2020). Accelerated epigenetic aging in adolescents from low-income households is associated with altered development of brain structures. *Metabolic Brain Disease*, *35*(8), 1287–1298. https://doi.org/10.1007/s11011-020-00589-0 | South Africa | Non-standard use |
| Nkomo, P., Mathee, A., Naicker, N., Galpin, J., Richter, L. M., & Norris, S. A. (2017). The association between elevated blood lead levels and violent behavior during late adolescence: The South African Birth to Twenty Plus cohort. *Environment International*, *109*, 136–145. https://doi.org/https://doi.org/10.1016/j.envint.2017.09.004 | South Africa | ASEBA data not reported |
| Nkomo, P., Naicker, N., Mathee, A., Galpin, J., Richter, L. M., & Norris, S. A. (2018). The association between environmental lead exposure with aggressive behavior, and dimensionality of direct and indirect aggression during mid-adolescence: Birth to Twenty Plus cohort. *Science of the Total Environment*, *612*, 472–479. https://doi.org/https://doi.org/10.1016/j.scitotenv.2017.08.138 | South Africa | Non-standard use |
| Nyamukapa, C. A., Gregson, S., Lopman, B., Saito, S., Watts, H. J., Monasch, R., & Jukes, M. C. H. (2008). HIV-Associated orphanhood and children's psychosocial distress: Theoretical framework tested with data from Zimbabwe. *American Journal of Public Health*, *98*(1), 133–141. https://doi.org/10.2105/AJPH.2007.116038 | Zimbabwe | Non-standard use |
| Reardon, C., George, G., Mucheuki, C., Govender, K., & Quinlan, T. (2015). Psychosocial and health risk outcomes among orphans and non-orphans in mixed households in KwaZulu-Natal, South Africa. *African Journal of AIDS Research*, *14*(4), 323–331. https://doi.org/10.2989/16085906.2015.1095774 | South Africa | Non-standard use |
| Spence, S. H., Donovan, C. L., March, S., Gamble, A., Anderson, R. E., Prosser, S., & Kenardy, J. (2011). A randomized controlled trial of online versus clinic-based CBT for adolescent anxiety. *Journal of Consulting and Clinical Psychology*, *79*(5), 629–642. https://doi.org/10.1037/a0024512 | South Africa (Australia) | No stratified data for South Africa participants |

| | | |
|---|---|---|
| Toska, E., Cluver, L., Orkin, M., Bains, A., Sherr, L., Berezin, M., & Gulaid, L. (2019). Screening and supporting through schools: educational experiences and needs of adolescents living with HIV in a South African cohort. *BMC Public Health*, *19*(1), N.PAG-N.PAG. https://doi.org/10.1186/s12889-019-6580-0 | South Africa | Non-standard use |
| Trude, A. C. B., Richter, L. M., Behrman, J. R., Stein, A. D., Menezes, A. M. B., & Black, M. M. (2021). Effects of responsive caregiving and learning opportunities during pre-school ages on the association of early adversities and adolescent human capital: an analysis of birth cohorts in two middle-income countries. *The Lancet Child & Adolescent Health*, *5*(1), 37–46. https://doi.org/https://doi.org/10.1016/S2352-4642(20)30309-6 | South Africa | Non-standard use |
| Waller, R., Gardner, F., & Cluver, L. (2014). Shared and unique predictors of antisocial and substance use behavior among a nationally representative sample of South African youth. *Aggression & Violent Behavior*, *19*(6), 629–636. https://doi.org/10.1016/j.avb.2014.09.002 | South Africa | Non-standard use |
| West Africa (*n* = 4) | | |
| Bishop, S. A., Owoloko, E. A., Okagbue, H. I., Oguntunde, P. E., Odetunmibi, O. A., & Opanuga, A. A. (2017). Survey datasets on the externalizing behaviors of primary school pupils and secondary school students in some selected schools in Ogun State, Nigeria. *Data in Brief*, *13*, 469–479. https://doi.org/10.1016/j.dib.2017.06.025 | Nigeria | Tool underwent extensive adaptation (e.g., with new items) |
| Bishop, S. A., Okagbue, H. I., & Odukoya, J. A. (2020). Statistical analysis of childhood and early adolescent externalizing behaviors in a middle low income country. *Heliyon, 6*(2), e03377. doi:10.1016/j.heliyon.2020.e03377 | Nigeria | Tool underwent extensive adaptation (e.g., with new items) |
| Bolton, S., McDonald, D., Curtis, E., Kelly, S., & Gallagher, L. (2014). Autism in a recently arrived immigrant population. *European Journal of Pediatrics*, *173*(3), 337–343. https://doi.org/10.1007/s00431-013-2149-6 | Nigeria and Congo | ASEBA data not reported |
| Roche, K. M., Bingenheimer, J. B., & Ghazarian, S. R. (2016). The dynamic interdependence between family support and depressive symptoms among adolescents in Ghana. *International Journal of Public Health*, *61*(4), 487–494. https://doi.org/10.1007/s00038-015-0781-9 | Ghana | Non-standard use |
| Central Africa (*n* = 2) | | |
| See Bolton et al. (2014) | Congo | |
| Ibinga, E., Ngoungou, E. B., Olliac, B., Hounsossou, C. H., Dalmay, F., Mouangue, G., Ategbo, S. J., Preux, P.-M., & Druet-Cabanac, M. (2015). Impact of epilepsy on children and parents in Gabon. *Epilepsy & Behavior*, *44*, 110–116. https://doi.org/10.1016/j.yebeh.2014.12.035 | Gabon | Non-standard use |
| Country Unknown (*n* = 20) | | |
| Benenson, J. F., Sinclair, N., & Dolenszky, E. (2006). Children's and adolescents' expectations of aggressive responses to provocation: Females predict more hostile reactions in compatible dyadic relationships. *Social Development*, *15*(1), 65–81. https://doi.org/10.1111/j.1467-9507.2006.00330.x | | Country of origin in Africa not specified |

| | |
|---|---|
| Cipolletta, S., Spina, G., & Spoto, A. (2018). Psychosocial functioning, self-image, and quality of life in children and adolescents with neurofibromatosis type 1. *Child: Care, Health & Development*, *44*(2), 260–268. https://doi.org/10.1111/cch.12496 | Country of origin in Africa not specified |
| de la Osa, N., Granero, R., Trepat, E., Domenech, J., & Ezpeleta, L. (2016). The discriminative capacity of CBCL/1½-5-DSM5 scales to identify disruptive and internalizing disorders in preschool children. *European Child & Adolescent Psychiatry*, *25*(1), 17–23. https://doi.org/10.1007/s00787-015-0694-4 | Country of origin in Africa not specified |
| Deater-Deckard, K., & Petrill, S. A. (2004). Parent–child dyadic mutuality and child behavior problems: an investigation of gene–environment processes. *Journal of Child Psychology & Psychiatry*, *45*(6), 1171–1179. https://doi.org/10.1111/j.1469-7610.2004.00309.x | Country of origin in Africa not specified |
| Derluyn, I., & Broekaert, E. (2007). Different perspectives on emotional and behavioural problems in unaccompanied refugee children and adolescents. *Ethnicity & Health*, *12*(2), 141–162. https://doi.org/10.1080/13557850601002296 | Country of origin in Africa not specified |
| Doom, J. R., Georgieff, M. K., & Gunnar, M. R. (2015). Institutional care and iron deficiency increase ADHD symptomology and lower IQ 2.5-5 years post-adoption. *Developmental Science*, *18*(3), 484–494. https://doi.org/10.1111/desc.12223 | Country of origin in Africa not specified |
| Heikkilä, A.-R., Elovainio, M., Raaska, H., Matomäki, J., Sinkkonen, J., & Lapinleimu, H. (2021). Intestinal parasites may be associated with later behavioral problems in internationally adopted children. *PLoS One*, *16*(1), e0245786. https://doi.org/https://doi.org/10.1371/journal.pone.0245786 | Country of origin in Africa not specified |
| Jackson, M. I., Kiernan, K., & McLanahan, S. (2012). Immigrant-native differences in child health: Does maternal education narrow or widen the gap? *Child Development*, *83*(5), 1501–1509. https://doi.org/10.1111/j.1467-8624.2012.01811.x | Country of origin in Africa not specified |
| Leventhal, T., & Shuey, E. A. (2014). Neighborhood context and immigrant young children's development. *Developmental Psychology*, *50*(6), 1771–1787. https://doi.org/10.1037/a0036424 | Country of origin in Africa not specified |
| Löfholm, C. A., Olsson, T., Sundell, K., & Hansson, K. (2009). Multisystemic therapy with conduct-disordered young people: Stability of treatment outcomes two years after intake. *Evidence & Policy: A Journal of Research, Debate and Practice*, *5*(4), 373–397. https://doi.org/https://doi.org/10.1332/174426409x478752 | Country of origin in Africa not specified |
| Meir, Y., Slone, M., & Lavi, I. (2012). Children of illegal migrant workers: Life circumstances and mental health. *Children & Youth Services Review*, *34*(8), 1546–1552. https://doi.org/10.1016/j.childyouth.2012.04.008 | Country of origin in Africa not specified |
| Meir, Y., Slone, M., & Levis, M. (2014). A randomized controlled study of a group intervention program to enhance mental health of children of illegal migrant workers. *Child & Youth Care Forum*, *43*(2), 165–180. https://doi.org/10.1007/s10566-013-9237-7 | Country of origin in Africa not specified |

| | |
|---|---|
| Pace, C. S., Di Folco, S., & Guerriero, V. (2018). Late-adoptions in adolescence: Can attachment and emotion regulation influence behaviour problems? A controlled study using a moderation approach. *Clinical Psychology & Psychotherapy*, *25*(2), 250–262. https://doi.org/10.1002/cpp.2158 | Country of origin in Africa not specified |
| Pearl, E. S. (2008). Parent-child interaction therapy with an immigrant family exposed to domestic violence. *Clinical Case Studies*, *7*(1), 25–41. https://doi.org/10.1177/1534650107300939 | Country of origin in Africa not specified; case study |
| Scheper, F. Y., Abrahamse, M. E., Jonkman, C. S., Schuengel, C., Lindauer, R. J. L., De Vries, A. L. C., Doreleijers, T. A. H., & Jansen, L. M. C. (2016). Inhibited attachment behaviour and disinhibited social engagement behaviour as relevant concepts in referred home reared children. *Child: Care, Health and Development*, *42*(4), 544–552. https://doi.org/https://doi.org/10.1111/cch.12319 | Country of origin in Africa not specified |
| Stellern, S., Esposito, E., Mliner, S., Pears, K., & Gunnar, M. (2014). Increased freezing and decreased positive affect in postinstitutionalized children. *Journal of Child Psychology & Psychiatry*, *55*(1), 88–95. https://doi.org/10.1111/jcpp.12123 | Country of origin in Africa not specified |
| Sundell, K., Hansson, K., Löfholm, C. A., Olsson, T., Gustle, L.-H., & Kadesjö, C. (2008). The transportability of multisystemic therapy to Sweden: Short-term results from a randomized trial of conduct-disordered youths. *Journal of Family Psychology*, *22*(4), 550–560. https://doi.org/10.1037/a0012790 | Country of origin in Africa not specified |
| van Ee, E., Kleber, R. J., & Mooren, T. T. M. (2012). War trauma lingers on: Associations between maternal posttraumatic stress disorder, parent-child interaction, and child development. *Infant Mental Health Journal*, *33*(5), 459–468. https://doi.org/10.1002/imhj.21324 | Country of origin in Africa not specified |
| Villabø, M., Gere, M., Torgersen, S., March, J., & Kendall, P. (2012). Diagnostic efficiency of the child and parent versions of the multidimensional anxiety scale for children. *Journal of Clinical Child & Adolescent Psychology*, *41*(1), 75–85. https://doi.org/10.1080/15374416.2012.632350 | Country of origin in Africa not specified |
| Webb, H. J., Thomas, R., McGregor, L., Avdagic, E., & Zimmer-Gembeck, M. J. (2017). An evaluation of parent–child interaction therapy with and without motivational enhancement to reduce attrition. *Journal of Clinical Child & Adolescent Psychology*, *46*(4), 537–550. https://doi.org/10.1080/15374416.2016.1247357 | Country of origin in Africa not specified |

**Appendix 1**

*Journal articles with ASEBA forms, a sub-Saharan African sample, but no psychometric properties (n = 87)*

**South Africa (*n* = 41)**

Asanbe, C., Moleko, A.-G., Visser, M., Thomas, A., Makwakwa, C., Salgado, W., & Tesnakis, A. (2016). Parental HIV/AIDS and psychological health of younger children in South Africa. *Journal of Child & Adolescent Mental Health*, *28*(2), 175–185. https://doi.org/10.2989/17280583.2016.1216853

Barbarin, O. A., Richter, L., & deWet, T. (2001). Exposure to violence, coping resources, and psychological adjustment of South African children. *The American Journal of Orthopsychiatry*, *71*(1), 16–25. https://doi.org/10.1037/0002-9432.71.1.16

Bell, C. C., Bhana, A., Petersen, I., McKay, M. M., Gibbons, R., Bannon, W., & Amatya, A. (2008). Building protective factors to offset sexually risky behaviors among black youths: a randomized control trial. *Journal of the National Medical Association*, *100*(8), 936–944. https://doi.org/10.1016/s0027-9684(15)31408-5

Bozicevic, L., De Pascalis, L., Schuitmaker, N., Tomlinson, M., Cooper, P. J., & Murray, L. (2016). Longitudinal association between child emotion regulation and aggression, and the role of parenting: A comparison of three cultures. *Psychopathology*, *49*(4), 228–235. https://doi.org/10.1159/000447747

Cluver, C. A., Charles, W., van der Merwe, C., Bezuidenhout, H., Nel, D., Groenewald, C., Brink, L., Hesselman, S., Bergman, L., & Odendaal, H. (2019). The association of prenatal alcohol exposure on the cognitive abilities and behaviour profiles of 4-year-old children: a prospective cohort study. *Bjog*, *126*(13), 1588–1597. https://doi.org/10.1111/1471-0528.15947

Cluver, L. D., Meinck, F., Steinert, J. I., Shenderovich, Y., Doubt, J., Romero, R. H., Lombard, C. J., Redfern, A., Ward, C. L., Tsoanyane, S., Nzima, D., Sibanda, N., Wittesaele, C., De Stone, S., Boyes, M. E., Catanho, R., Lachman, J. M., Salah, N., Nocuza, M., & Gardner, F. (2018). Parenting for lifelong health: A pragmatic cluster randomised controlled trial of a non-commercialised parenting programme for adolescents and their families in South Africa. *BMJ Global Health*, *3*(1). https://doi.org/10.1136/bmjgh-2017-000539

Cluver, L. D., Gardner, F., & Operario, D. (2008). Effects of stigma on the mental health of adolescents orphaned by AIDS. *Journal of Adolescent Health*, *42*(4), 410–417. https://doi.org/10.1016/j.jadohealth.2007.09.022

Cluver, L. D., Orkin, F. M., Meinck, F., Boyes, M. E., Yakubovich, A. R., & Sherr, L. (2016). Can Social Protection Improve Sustainable Development Goals for Adolescent Health? *PLoS One*, *11*(10), 1–20. https://doi.org/10.1371/journal.pone.0164808

Cluver, L. D., Orkin, F. M., Meinck, F., Boyes, M. E., & Sherr, L. (2016). Structural drivers and social protection: mechanisms of HIV risk and HIV prevention for South African adolescents. *Journal of the International AIDS Society*, *19*(1), 20646. https://doi.org/10.7448/ias.19.1.20646

Cluver, L., Gardner, F., & Operario, D. (2007). Psychological distress amongst AIDS-orphaned children in urban South Africa. *Journal of Child Psychology & Psychiatry*, *48*(8), 755–763. https://doi.org/10.1111/j.1469-7610.2007.01757.x

Cluver, L., Gardner, F., & Operario, D. (2009). Poverty and psychological health among AIDS-orphaned children in Cape Town, South Africa. *AIDS Care*, *21*(6), 732–741. https://doi.org/10.1080/09540120802511885

Cluver, L., Meinck, F., Shenderovich, Y., Ward, C. L., Romero, R. H., Redfern, A., Lombard, C., Doubt, J., Steinert, J., Catanho, R., Wittesaele, C., De Stone, S., Salah, N., Mpimpilashe, P., Lachman, J., Loening, H., Gardner, F., Blanc, D., Nocuza, M., & Lechowicz, M. (2016). A parenting programme to prevent abuse of adolescents in South Africa: Study protocol for a randomised controlled trial. *Trials*, *17*(1), 328. https://doi.org/10.1186/s13063-016-1452-8

Cluver, L., Operario, D., & Gardner, F. (2009). Parental illness, caregiving factors and psychological distress among children orphaned by acquired immune deficiency syndrome (AIDS) in South Africa. *Vulnerable Children and Youth Studies*, *4*(3), 185–198. https://doi.org/10.1080/17450120902730196

[a]Cortina, M. A., Stein, A., Kahn, K., Hlungwani, T. M., Holmes, E. A., & Fazel, M. (2016). Cognitive styles and psychological functioning in rural South African school students: Understanding influences for risk and resilience in the face of chronic adversity. *Journal of Adolescence*, *49*, 38–46. https://doi.org/10.1016/j.adolescence.2016.01.010

De Stone, S., F. Meinck, L. Sherr, et al. (2016). Factors associated with good and harsh parenting of pre-adolescents and adolescents in Southern Africa, *Innocenti Working Paper* No. 2016-20, UNICEF Office of Research, Florence.

Dollman, A. K., Figaji, A. A., & Schrieff-Elson, L. E. (2017). Academic and Behavioral Outcomes in School-Age South African Children Following Severe Traumatic Brain Injury. *Frontiers in Neuroanatomy*, *11*. https://doi.org/10.3389/fnana.2017.00121

Donald, K. A. M., Mathema, H., Thomas, K. G. F., & Wilmshurst, J. M. (2011). Intellectual and behavioral functioning in a South African cohort of boys with Duchenne muscular dystrophy. *Journal of Child Neurology*, *26*(8), 963–969. https://doi.org/10.1177/0883073811399149

Ebersöhn, L., Eloff, I., Finestone, M., Grobler, A., & Moen, M. (2015). Telling stories and adding scores: Measuring resilience in young children affected by maternal HIV and AIDS. *African Journal of AIDS Research*, *14*(3), 219–227. https://doi.org/10.2989/16085906.2015.1052822

Eloff, I., Finestone, M., Makin, J. D., Boeving-Allen, A., Visser, M., Ebersöhn, L., Ferreira, R., Sikkema, K. J., Briggs-Gowan, M. J., & Forsyth, B. W. C. (2014). A randomized clinical trial of an intervention to promote resilience in young children of HIV-positive mothers in South Africa. *Aids*, *28*(SUPPL. 3), S347–S357. https://doi.org/10.1097/QAD.0000000000000335

Eloff, I., Finestone, M., & Forsyth, B. (2016). HIV/AIDS Infected Mothers' Experience of a Group Intervention to Enhance Their Children's Behavior. *South African Journal of Education*, *36*(2). http://dx.doi.org/10.15700/saje.v36n2a1285

Garman, E. C., Cois, A., Tomlinson, M., Rotheram-Borus, M. J., & Lund, C. (2019). Course of perinatal depressive symptoms among South African women: associations with child outcomes at 18 and 36 months. *Social Psychiatry and Psychiatric Epidemiology*, 1–13. https://doi.org/10.1007/s00127-019-01665-2

Hoare, J., Phillips, N., Brittain, K., Myer, L., Zar, H. J., & Stein, D. J. (2019). Mental Health and Functional Competence in the Cape Town Adolescent Antiretroviral Cohort. *J Acquir Immune Defic Syndr*, *81*(4), e109–e116. https://doi.org/10.1097/qai.0000000000002068

Hoare, J., Fouche, J.-P., Phillips, N., Joska, J. A., Myer, L., Zar, H. J., & Stein, D. J. (2018). Structural brain changes in perinatally HIV-infected young adolescents in South Africa. *AIDS*, *32*(18), 2707–2718. https://doi.org/10.1097/QAD.0000000000002024

Hoare, J., Phillips, N., Joska, J. A., Paul, R., Donald, K. A., Stein, D. J., & Thomas, K. G. F. (2016). Applying the HIV-associated neurocognitive disorder diagnostic criteria to HIV-infected youth. *Neurology*, *87*(1), 86–93. https://doi.org/10.1212/WNL.0000000000002669

Lanesman, T. H., & Schrieff, L. E. (2020). Implementation of an attention training programme with a sample of children who have sustained traumatic brain injuries in South Africa: A pilot study. *Neuropsychological Rehabilitation*, 1–29. https://doi.org/10.1080/09602011.2020.1782233

Louw, K.-A., Ipser, J., Phillips, N., & Hoare, J. (2016). Correlates of emotional and behavioural problems in children with perinatally acquired HIV in Cape Town, South Africa. *AIDS Care*, *28*(7), 842–850. https://doi.org/10.1080/09540121.2016.1140892

Naicker, N., Richter, L., Mathee, A., Becker, P., & Norris, S. A. (2012). Environmental lead exposure and socio-behavioural adjustment in the early teens: the birth to twenty cohort. *Science of the Total Environment*, *414*, 120–125. https://doi.org/10.1016/j.scitotenv.2011.11.013

Nothling, J., Martin, C. L., Laughton, B., Cotton, M. F., & Seedat, S. (2013). Maternal post-traumatic stress disorder, depression and alcohol dependence and child behaviour outcomes in mother-child dyads infected with HIV: a longitudinal study. *BMJ Open*, *3*(12), e003638. https://doi.org/10.1136/bmjopen-2013-003638

[b]Phillips, N., Thomas, K. G. F., Mtukushe, B., Myer, L., Zar, H. J., Stein, D. J., & Hoare, J. (2021). Youth perinatal HIV-associated neurocognitive disorders: association with functional impairment. *AIDS Care*, 1–5. https://doi.org/10.1080/09540121.2021.1891191

Rochat, T. J., Houle, B., Stein, A., Mitchell, J., & Bland, R. M. (2019). Maternal alcohol use and children's emotional and cognitive outcomes in rural South Africa. *SAMJ: South African Medical Journal*, *109*(7), 526–534. https://doi.org/10.7196/samj.2019.v109i7.13120

Rochat, T., Stein, A., Cortina-Borja, M., Tanser, F., & Bland, R. M. (2017). The Amagugu intervention to increase disclosure of maternal HIV to HIV-uninfected primary-school aged children in Southern Africa: A randomised controlled trial. *Lancet HIV*, *4*(12), e566–e576. https://doi.org/10.1016/s2352-3018(17)30133-9

Rochat, T. J., Arteche, A. X., Stein, A., Mkwanazi, N., & Bland, R. M. (2014). Maternal HIV disclosure to young HIV-uninfected children: an evaluation of a family-centred intervention in South Africa. *AIDS (London, England)*, *28 Suppl 3*, S331-41. https://doi.org/10.1097/QAD.0000000000000333

Rochat, T. J., Houle, B., Stein, A., Pearson, R. M., & Bland, R. M. (2018). Prevalence and risk factors for child mental disorders in a population-based cohort of HIV-exposed and unexposed African children aged 7–11 years. *European Child & Adolescent Psychiatry*, *27*(12), 1607–1620. https://doi.org/10.1007/s00787-018-1146-8

Rossouw, J., Yadin, E., Alexander, D., & Seedat, S. (2018). Prolonged exposure therapy and supportive counselling for posttraumatic stress disorder in adolescents in a community-based sample, including experiences of stakeholders: study protocol for a comparative randomized controlled trial using task-shifting. *BMC Psychiatry*, *18*(1), N.PAG-N.PAG. https://doi.org/10.1186/s12888-018-1873-x

Rotheram-Borus, M. J., Arfer, K. B., Christodoulou, J., Comulada, W. S., Stewart, J., Tubert, J. E., & Tomlinson, M. (2019). The association of maternal alcohol use and paraprofessional home visiting with children's health: A randomized controlled trial. *Journal of Consulting and Clinical Psychology*, *87*(6), 551–562. https://doi.org/10.1037/ccp0000408

Rotheram-Fuller, E. J., Tomlinson, M., Scheffler, A., Weichle, T. W., Hayati Rezvan, P., Comulada, W. S., & Rotheram-Borus, M. J. (2018). Maternal patterns of antenatal and postnatal depressed mood and the impact on child health at 3-years postpartum. *Journal of Consulting and Clinical Psychology*, *86*(3), 218–230. https://doi.org/10.1037/ccp0000281

Sabet, F., Richter, L. M., Ramchandani, P. G., Stein, A., Quigley, M. A., & Norris, S. A. (2009). Low birthweight and subsequent emotional and behavioural outcomes in 12-year-old children in Soweto, South Africa: Findings from Birth to Twenty. *International Journal of Epidemiology*, *38*(4), 944–954. https://doi.org/10.1093/ije/dyp204

St Clair, M. C., Skeen, S., Marlow, M., & Tomlinson, M. (2019). Relationships between concurrent language ability and mental health outcomes in a South African sample of 13-year-olds. *PLoS One*, *14*(9), e0221242. https://doi.org/10.1371/journal.pone.0221242

van Dyk, J., Ramanjam, V., Church, P., Koren, G., & Donald, K. (2014). Maternal methamphetamine use in pregnancy and long-term neurodevelopmental and behavioral deficits in children. *Journal of Population Therapeutics and Clinical Pharmacology*, *21*(2), e185–e196.

Wait, J. W. V. (2002). Tuberculosis Meningitis and Attention Deficit Hyperactivity Disorder in Children. *Journal of Tropical Pediatrics*, *48*(5), 294–299. https://doi.org/10.1093/tropej/48.5.294

Wait, J. W., & Schoeman, J. F. (2010). Behaviour Profiles After Tuberculous Meningitis. *Journal of Tropical Pediatrics*, *56*(3), 166–171. https://doi.org/10.1093/tropej/fmp080

[a]Internal consistency analyses were conducted but not reported.
[b]Psychometric analyses were performed but they were not conventional.

**Kenya (*n* = 18)**

Denckla, C. A., Ndetei, D. M., Mutiso, V. N., Musyimi, C. W., Musau, A. M., Nandoya, E. S., Anderson, K. K., Milanovic, S., Henderson, D., & McKenzie, K. (2017). Psychometric properties of the Ndetei-Othieno-Kathuku (NOK) Scale: A mental health assessment tool for an African setting. *J Child Adolesc Ment Health*, *29*(1), 39–49. https://doi.org/10.2989/17280583.2017.1310729

Di Giunta, L., Rothenberg, W. A., Lunetti, C., Lansford, J. E., Pastorelli, C., Eisenberg, N., Thartori, E., Basili, E., Favini, A., & Yotanyamaneewong, S. (2020). Longitudinal associations between mothers' and fathers' anger/irritability expressiveness, harsh parenting, and adolescents' socioemotional functioning in nine countries. *Developmental Psychology*, *56*(3), 458. https://doi.org/10.1037/dev0000849

Dodge, K. A., Malone, P. S., Lansford, J. E., Sorbring, E., Skinner, A. T., Tapanya, S., Uribe Tiradod, L. M., Zelli, A., Alampay, L. P., Al-Hassan, S. M., Bacchini, D., Bombi, A. S., Bornstein, M. H., Chang, L., Deater-Deckard, K., Di Giunta, L., Oburu, P., & Pastorelli, C. (2015). Hostile attributional bias and aggressive behavior in global context. *PNAS Proceedings of the National Academy of Sciences of the United States of America*, *112*(30), 9310–9315. https://doi.org/10.1073/pnas.1418572112

Karanja, S. W., Kiburi, S. K., Kang'ethe, R., & Othieno, C. J. (2021). Emotional and behavioral problems in children with epilepsy attending the pediatric neurology clinic at a referral hospital in Kenya. *Epilepsy Behav*, *114*(Pt A), 107477. https://doi.org/10.1016/j.yebeh.2020.107477

Kariuki, S. M., Abubakar, A., Kombe, M., Kazungu, M., Odhiambo, R., Stein, A., & Newton, C. R. J. C. (2017). Burden, risk factors, and comorbidities of behavioural and emotional problems in Kenyan children: a population-based study. *The Lancet Psychiatry*, *4*(2), 136–145. https://doi.org/10.1016/S2215-0366(16)30403-5

Kariuki, S. M., Abubakar, A., Kombe, M., Kazungu, M., Odhiambo, R., Stein, A., & Newton, C. R. J. C. (2018). Prevalence, risk factors and behavioural and emotional comorbidity of acute seizures in young Kenyan children: a population-based study. *BMC Medicine*, *16*(1), 35. https://doi.org/10.1186/s12916-018-1021-y

Lansford, J. E., Godwin, J., Al-Hassan, S. M., Bacchini, D., Bornstein, M. H., Chang, L., Chen, B.-B., Deater-Deckard, K., Di Giunta, L., Dodge, K. A., Malone, P. S., Oburu, P., Pastorelli, C., Skinner, A. T., Sorbring, E., Steinberg, L., Tapanya, S., Alampay, L. P., Uribe Tirado, L. M., & Zelli, A. (2018). Longitudinal associations between parenting and youth adjustment in twelve cultural groups: Cultural normativeness of parenting as a moderator. *Developmental Psychology*, *54*(2), 362–377. https://doi.org/10.1037/dev0000416

Lansford, J. E., Sharma, C., Malone, P. S., Woodlief, D., Dodge, K. A., Oburu, P., Pastorelli, C., Skinner, A. T., Sorbring, E., Tapanya, S., Tirado, L. M. U., Zelli, A., Al-Hassan, S. M., Alampay, L. P., Bacchini, D., Bombi, A. S., Bornstein, M. H., Chang, L., Deater-Deckard, K., & Di Giunta, L. (2014). Corporal punishment, maternal warmth, and child adjustment: A longitudinal study in eight countries. *Journal of Clinical Child and Adolescent Psychology*, *43*(4), 670–685. https://doi.org/10.1080/15374416.2014.893518

Magai, D. N., & Koot, H. M. (2019). Quality of life in children and adolescents in Central Kenya: associations with emotional and behavioral problems. *Quality of Life Research*, *28*(5), 1271–1279. https://doi.org/10.1007/s11136-019-02099-8

Mutiso, V., Musyimi, C., Tele, A., Gitonga, I., & Ndetei, D. (2020). Feasibility study on the mhGAP-IG as a tool to enhance parental awareness of symptoms of mental disorders in lower primary (6-10 year old) school-going children: Towards inclusive child mental health services in a Kenyan setting. *Early Interv Psychiatry*. https://doi.org/10.1111/eip.12963

Mutiso, V., Tele, A., Musyimi, C., Gitonga, I., Musau, A., & Ndetei, D. (2018). Effectiveness of life skills education and psychoeducation on emotional and behavioral problems among adolescents in institutional care in Kenya: a longitudinal study. *Child and Adolescent Mental Health*, *23*(4), 351–358. https://doi.org/10.1111/camh.12232

Mutiso, V. N., Musyimi, C. W., Tele, A., & Ndetei, D. M. (2017). Epidemiological patterns and correlates of mental disorders among orphans and vulnerable children under institutional care. *Social Psychiatry and Psychiatric Epidemiology*, *52*(1), 65–75. https://doi.org/10.1007/s00127-016-1291-7

Ndetei, D. M., Mutiso, V., Gitonga, I., Agudile, E., Tele, A., Birech, L., Musyimi, C., & McKenzie, K. (2019). World Health Organization life-skills training is efficacious in reducing youth self-report scores in primary school going children in Kenya. *Early Intervention in Psychiatry*, *13*(5), 1146–1154. https://doi.org/10.1111/eip.12745

Ndetei, D., Mutiso, V., Maraj, A., Anderson, K., Musyimi, C., Musau, A., Tele, A., Gitonga, I., & McKenzie, K. (2019). Towards Understanding the Relationship Between Psychosocial Factors and Ego Resilience Among Primary School Children in a Kenyan Setting: A Pilot Feasibility Study. *Community Mental Health Journal*, *55*(6), 1038–1046. https://doi.org/10.1007/s10597-019-00425-5

Ndetei, D., Mutiso, V., Musyimi, C., Mokaya, A., Anderson, K., McKenzie, K., Musau, A., Ndetei, D. M., Mokaya, A. G., & Anderson, K. K. (2016). The prevalence of mental disorders among upper primary school children in Kenya. *Social Psychiatry & Psychiatric Epidemiology*, *51*(1), 63–71. https://doi.org/10.1007/s00127-015-1132-0

Rothenberg, W. A., Lansford, J. E., Al-Hassan, S. M., Bacchini, D., Bornstein, M. H., Chang, L., Deater-Deckard, K., Di Giunta, L., Dodge, K. A., & Malone, P. S. (2020). Examining effects of parent warmth and control on internalizing behavior clusters from age 8 to 12 in 12 cultural groups in nine countries. *Journal of Child Psychology and Psychiatry*, *61*(4), 436–446. https://doi.org/10.1111/jcpp.13138

Schenck-Fontaine, A., Lansford, J. E., Skinner, A. T., Deater-Deckard, K., Di Giunta, L., Dodge, K. A., Oburu, P., Pastorelli, C., Sorbring, E., & Steinberg, L. (2020). Associations between perceived material deprivation, parents' discipline practices, and children's behavior problems: An international perspective. *Child Development*, *91*(1), 307–326. https://doi.org/10.1111/cdev.13151

Weisz, J. R., Sigman, M., Weiss, B., & Mosk, J. (1993). Parent reports of behavioral and emotional problems among children in Kenya, Thailand, and the United States. *Child Dev*, *64*(1), 98–109. https://doi.org/10.1111/j.1467-8624.1993.tb02897.x

**Uganda (*n* = 13)**

Bangirana, P., Allebeck, P., Boivin, M. J., John, C. C., Page, C., Ehnvall, A., & Musisi, S. (2011). Cognition, behaviour and academic skills after cognitive rehabilitation in Ugandan children surviving severe malaria: a randomised trial. *BMC Neurology*, *11*(1), 96. https://doi.org/10.1186/1471-2377-11-96

Bangirana, P., Giordani, B., John, C. C., Page, C., Opoka, R. O., & Boivin, M. J. (2009). Immediate Neuropsychological and Behavioral Benefits of Computerized Cognitive Rehabilitation in Ugandan Pediatric Cerebral Malaria Survivors. *Journal of Developmental & Behavioral Pediatrics*, *30*(4), 310–318. https://doi.org/10.1097/DBP.0b013e3181b0f01b

Bangirana, P., Musisi, S., Boivin, M. J., Ehnvall, A., John, C. C., Bergemann, T. L., & Allebeck, P. (2011). Malaria with neurological involvement in Ugandan children: effect on cognitive ability, academic achievement and behaviour. *Malaria Journal*, *10*(1), 334. https://doi.org/10.1186/1475-2875-10-334

Boivin, M. J., Sikorskii, A., Nakasujja, N., Ruisenor-Escudero, H., Familiar-Lopez, I., Opoka, R. O., & Giordani, B. (2019). Evaluating Immunopathogenic Biomarkers During Severe Malaria Illness as Modifiers of the Neuropsychologic Benefits of Computer Cognitive Games Rehabilitation in Ugandan Children. *Pediatr Infect Dis J*, *38*(8), 840–848. https://doi.org/10.1097/inf.0000000000002367

Boivin, M. J., Bangirana, P., Nakasujja, N., Page, C. F., Shohet, C., Givon, D., Bass, J. K., Opoka, R. O., & Klein, P. S. (2013). A Year-Long Caregiver Training Program Improves Cognition in Preschool Ugandan Children with Human Immunodeficiency Virus. *The Journal of Pediatrics*, *163*(5), 1409-1416.e5. https://doi.org/10.1016/j.jpeds.2013.06.055

Boivin, M. J., Bangirana, P., Nakasujja, N., Page, C. F., Shohet, C., Givon, D., Bass, J. K., Opoka, R. O., & Klein, P. S. (2013). A Year-long Caregiver Training Program to Improve Neurocognition in Preschool Ugandan HIV-exposed Children. *Journal of Developmental & Behavioral Pediatrics*, *34*(4), 269–278. https://doi.org/10.1097/DBP.0b013e318285fba9

Boivin, M. J., Nakasujja, N., Sikorskii, A., Opoka, R. O., & Giordani, B. (2016). A Randomized Controlled Trial to Evaluate if Computerized Cognitive Rehabilitation Improves Neurocognition in Ugandan Children with HIV. *AIDS Research and Human Retroviruses*, *32*(8), 743–755. https://doi.org/10.1089/aid.2016.0026

Boivin, M. J., Nakasujja, N., Sikorskii, A., Ruiseñor-Escudero, H., Familiar-Lopez, I., Walhof, K., van der Lugt, E. M., Opoka, R. O., & Giordani, B. (2019). Neuropsychological benefits of computerized cognitive rehabilitation training in Ugandan children surviving severe malaria: A randomized controlled trial. *Brain Research Bulletin*, *145*, 117–128. https://doi.org/10.1016/j.brainresbull.2018.03.002

Hickson, M. R., Conroy, A. L., Bangirana, P., Opoka, R. O., Idro, R., Ssenkusu, J. M., & John, C. C. (2019). Acute kidney injury in Ugandan children with severe malaria is associated with long-term behavioral problems. *PLoS One*, *14*(12), e0226405. https://doi.org/10.1371/journal.pone.0226405

Ojiambo, D., & Bratton, S. C. (2014). Effects of group activity play therapy on problem behaviors of preadolescent Ugandan orphans. *Journal of Counseling & Development*, *92*(3), 355–365. https://doi.org/10.1002/j.1556-6676.2014.00163.x

Okello, J., Nakimuli-Mpungu, E., Musisi, S., Broekaert, E., & Derluyn, I. (2013). War-related trauma exposure and multiple risk behaviors among school-going adolescents in Northern Uganda: the mediating role of depression symptoms. *J Affect Disord*, *151*(2), 715–721. https://doi.org/10.1016/j.jad.2013.07.030

Ssemata, A. S., Hickson, M., Ssenkusu, J. M., Cusick, S. E., Nakasujja, N., Opoka, R. O., Kroupina, M., Georgieff, M. K., Bangirana, P., & John, C. C. (2020). Delayed iron does not alter cognition or behavior among children with severe malaria and iron deficiency. *Pediatric Research*, *88*(3), 429–437. https://doi.org/10.1038/s41390-020-0957-8

Ssenkusu, J. M., Hodges, J. S., Opoka, R. O., Idro, R., Shapiro, E., John, C. C., & Bangirana, P. (2016). Long-term Behavioral Problems in Children With Severe Malaria. *Pediatrics*, *138*(5). https://doi.org/10.1542/peds.2016-1965

**Ethiopia (*n* = 4)**

Rescorla, L., Achenbach, T. M., Ivanova, M. Y., Dumenci, L., Almqvist, F., Bilenberg, N., Bird, H., Broberg, A., Dobrean, A., Döpfner, M., Erol, N., Forns, M., Hannesdottir, H., Kanbayashi, Y., Lambert, M. C., Leung, P., Minaei, A., Mulatu, M. S., Novik, T. S., … Verhulst, F. (2007). Epidemiological comparisons of problems and positive qualities reported by adolescents in 24 countries. *Journal of Consulting and Clinical Psychology*, *75*(2), 351–358. https://doi.org/10.1037/0022-006X.75.2.351

Rescorla, L., Achenbach, T., Ivanova, M. Y., Dumenci, L., Almqvist, F., Bilenberg, N., Bird, H., Chen, W., Dobrean, A., Döpfner, M., Erol, N., Fombonne, E., Fonseca, A., Frigerio, A., Grietens, H., Hannesdottir, H., Kanbayashi, Y., Lambert, M., Larsson, B., … Verhulst, F. (2007). Behavioral and emotional problems reported by parents of children ages 6 to 16 in 31 societies. *Journal of Emotional and Behavioral Disorders*, *15*(3), 130–142. https://doi.org/10.1177/10634266070150030101

Rescorla, L., Ivanova, M. Y., Achenbach, T. M., Begovac, I., Chahed, M., Drugli, M. B., Emerich, D. R., Fung, D. S. S., Haider, M., Hansson, K., Hewitt, N., Jaimes, S., Larsson, B., Maggiolini, A., Marković, J., Mitrović, D., Moreira, P., Oliveira, J. T., Olsson, M., … Zhang, E. Y. (2012). International epidemiology of child and adolescent psychopathology II: Integration and applications of dimensional findings from 44 societies. *Journal of the American Academy of Child & Adolescent Psychiatry*, *51*(12), 1273–1283. https://doi.org/10.1016/j.jaac.2012.09.012

Tadesse, A. W., Berhane Tsehay, Y., Girma Belaineh, B., & Alemu, Y. B. (2012). Behavioral and emotional problems among children aged 6–14 years on highly active antiretroviral therapy in Addis Ababa: A cross-sectional study. *AIDS Care*, *24*(11), 1359–1367. https://doi.org/10.1080/09540121.2011.650677

**Malawi (*n* = 4)**

Boivin, M. J., Gladstone, M. J., Vokhiwa, M., Birbeck, G. L., Magen, J. G., Page, C., Semrud-Clikeman, M., Kauye, F., & Taylor, T. E. (2011). Developmental outcomes in Malawian children with retinopathy-positive cerebral malaria. *Tropical Medicine & International Health*, *16*(3), 263–271. https://doi.org/10.1111/j.1365-3156.2010.02704.x

Boivin, M. J., Mohanty, A., Sikorskii, A., Vokhiwa, M., Magen, J. G., & Gladstone, M. (2019). Early and middle childhood developmental, cognitive, and psychiatric outcomes of Malawian children affected by retinopathy positive cerebral malaria. *Child Neuropsychology*, *25*(1), 81–102. https://doi.org/10.1080/09297049.2018.1451497

Boivin, M. J., Vokhiwa, M., Sikorskii, A., Magen, J. G., & Beare, N. A. V. (2014). Cerebral Malaria Retinopathy Predictors of Persisting Neurocognitive Outcomes in Malawian Children. *The Pediatric Infectious Disease Journal*, *33*(8), 821–824. https://doi.org/10.1097/INF.0000000000000296

Madhlopa, Y., Qin, J., & Chen, C. (2020). The relationships between child maltreatment and child behavior problems. Comparative study of Malawi and China. *Children and Youth Services Review*, *119*, 105533. https://doi.org/10.1016/j.childyouth.2020.105533

**Democratic Republic of the Congo (*n* = 1)**

Matonda-ma-Nzuzi, T., Mampunza Ma Miezi, S., Mpembi, M. N., Mvumbi, D. M., Aloni, M. N., Malendakana, F., Mpaka Mbeya, D., Lelo, G. M., & Charlier-Mikolajczak, D. (2018). Factors associated with behavioral problems and cognitive impairment in children with epilepsy of Kinshasa, Democratic Republic of the Congo. *Epilepsy & Behavior*, *78*, 78–83. https://doi.org/10.1016/j.yebeh.2017.08.030

**Ghana (*n* = 1)**

Kusi-Mensah, K., Donnir, G., Wemakor, S., Owusu-Antwi, R., & Omigbodun, O. (2019). Prevalence and patterns of mental disorders among primary school age children in Ghana: correlates with academic achievement. *J Child Adolesc Ment Health*, *31*(3), 214–223. https://doi.org/10.2989/17280583.2019.1678477

**Tanzania (*n* = 1)**

Van de Wijer, L., Mchaile, D. N., de Mast, Q., Mmbaga, B. T., Rommelse, N. N. J., Duinmaijer, A., van der Ven, A. J. A. M., Schellekens, A. F. A., & Kinabo, G. D. (2019). Neuropsychiatric symptoms in Tanzanian HIV-infected children receiving long-term efavirenz treatment: a multicentre, cross-sectional, observational study. *The Lancet HIV*, *6*(4), e250–e258. https://doi.org/10.1016/S2352-3018(18)30329-1

**Immigrant samples (*n* = 4)**

Huemer, J., Karnik, N., Voelkl-Kernstock, S., Granditsch, E., Plattner, B., Friedrich, M., & Steiner, H. (2011). Psychopathology in African Unaccompanied Refugee Minors in Austria. *Child Psychiatry & Human Development*, *42*(3), 307–319. https://doi.org/10.1007/s10578-011-0219-4

Huemer, J., Völkl-Kernstock, S., Karnik, N., G. Denny, K., Granditsch, E., Mitterer, M., Humphreys, K., Plattner, B., Friedrich, M., Shaw, R. J., & Steiner, H. (2013). Personality and Psychopathology in African Unaccompanied Refugee Minors: Repression, Resilience and Vulnerability. *Child Psychiatry & Human Development*, *44*(1), 39–50. https://doi.org/10.1007/s10578-012-0308-z

Sourander, A. (1998). Behavior Problems and Traumatic Events of Unaccompanied Refugee Minors. *Child Abuse & Neglect*, *22*(7), 719–727. https://doi.org/https://doi.org/10.1016/S0145-2134(98)00053-2

Betancourt, T. S., Berent, J. M., Freeman, J., Frounfelker, R. L., Brennan, R. T., Abdi, S., Maalim, A., Abdi, A., Mishra, T., & Gautam, B. (2020). Family-based mental health promotion for Somali bantu and Bhutanese refugees: feasibility and acceptability trial. *Journal of Adolescent Health*, *66*(3), 336–344. https://doi.org/10.1016/j.jadohealth.2019.08.023
